# Defining the extracellular matrix in non-cartilage soft tissues in osteoarthritis – a systematic review

**DOI:** 10.1101/2023.08.31.23294625

**Authors:** Jolet Y. Mimpen, Iwan G. A. Raza, Sarah J. B. Snelling

## Abstract

**Objective:** Osteoarthritis (OA) is increasingly seen as a disease of global joint dysfunction, affecting not only cartilage but also the other joint tissues. Extracellular matrix (ECM) is a critical determinant of tissue mechanobiology, but ECM is poorly understood in osteoarthritic joint tissues beyond cartilage in human OA and animal models of OA. Therefore, we aimed to define the structural composition and architecture of non-cartilage soft joint tissue ECM in human OA, and to compare the ECM changes observed in humans to those seen in animal models of OA.

**Design:** A systematic search strategy, devised using relevant matrix, tissue, and disease nomenclature, was run through the MEDLINE, EMBASE, and Scopus databases. Demographic, clinical, and biological data were extracted from eligible studies. Bias analysis was performed.

**Results:** 142 studies were included, which covered capsule, ligaments, meniscus, skeletal muscle, synovium, and tendon in both humans and animals, and fat pad and intervertebral disc in humans only. Overall, included studies show that the expression of structural ECM components changes in disease within an ECM that becomes disorganised with increasing joint degeneration.

**Conclusions:** This systematic review consolidates existing knowledge of a poorly defined aspect of OA pathophysiology. Changes in ECM composition and architecture occur across soft joint tissues in OA, but most of these remain poorly defined due to the low number of studies and lack of healthy comparator groups. Further research to better understand the context within which cartilage is damaged in OA may enable a better understanding of OA and its potential treatments.

**Key messages:** *What is already known on this topic:*

- Extracellular matrix (ECM) is a critical determinant of tissue mechanobiology and cell behaviour, but it is poorly described in osteoarthritic joint tissues beyond cartilage.

*What this study adds:*

- Our study highlights the global nature of ECM dysregulation across the osteoarthritic joint. In addition, this study describes practical and methodological challenges that should be addressed to improve the contribution of future studies to define the role of ECM in non-cartilage soft tissues in osteoarthritis.

*How this study might affect research, practice or policy:*

- A better understanding of ECM changes and their underlying mechanisms throughout the osteoarthritic joint may assist with disease classification and patient stratification and also holds promise for the development of ECM-targeting treatments which could modify the pathogenic cell behaviour that may drive osteoarthritis progression.

## 1 Introduction

Osteoarthritis (OA) is the most common joint disease globally, affecting over 500 million people. OA is typically attributed to mechanically-driven joint damage and is characterised by articular cartilage degeneration and subchondral bone remodelling[1]. However, these tissues are not affected in isolation from the wider joint, with pathology in other soft joint tissues contributing to the symptoms and progression of OA[2, 3]. Damage to menisci and ligaments disrupts joint biomechanics, while inflammation, fibrosis, and distension of the synovium and joint capsule are associated with joint pain and stiffness[4–8]. Despite significant clinical need and substantial efforts to identify disease modifying OA drugs, there is no effective way of inhibiting or decelerating OA-related joint damage by targeting cartilage directly. Given the important role of other soft tissues in joint biomechanics and the release of pro-inflammatory and matrix-degrading mediators into the synovial fluid[9], understanding the whole joint context of OA might provide novel therapeutic strategies and prognostic markers.

Joint tissues are rich in extracellular matrix (ECM), a network of structural and regulatory macromolecules within which cells are embedded[10]. The role of ECM as a major determinant of the biophysical properties of a tissue has clear relevance in a disease such as OA[11, 12]. ECM not only provides structure to the tissue, but can also affect cell function through receptor engagement, mechanical cues, and the sequestration of growth factors and cytokines[13–16]. Significant crosstalk occurs between cells and matrix components, such that pathological ECM may exacerbate cellular dysfunction in disease[15, 17]. Therefore, ECM composition and architecture cannot be disregarded when attempting to understand OA pathophysiology. However, outside of cartilage, ECM remodelling in OA has received relatively little attention.

Studying OA in the clinical setting is challenging due to the slow and unpredictable nature of the course of the disease. In addition, clinical symptoms often appear late in the disease process, making it difficult to study its onset and early progression. Therefore, many animal models for OA have been developed and are used to overcome these issues and to facilitate the development and evaluation of new therapies and diagnostic tools[18]. However, since there is no single “gold standard” animal model that accurately reflects all aspects of human disease, a major challenge is selecting the “right” model for each study[19].

The main aim of this systematic review is to consolidate existing data describing ECM architecture and structural composition in the synovium, joint capsule, skeletal muscle, tendon, ligament, meniscus, intervertebral disc, and fat pad of osteoarthritic joints. Secondly, we aim to define the changes in ECM architecture and structural composition in these tissues in animal models of OA to address their ability to replicate human disease pathophysiology.

## 2 Methods

This review was conducted according to a protocol registered on the PROSPERO database [CRD42021231241] and guidelines set out in the Preferred Reporting Items for Systematic Reviews and Meta-Analyses (PRISMA) statement[20].

### Database and search strategy

The search strategy, written by JM and a medical librarian, can be found in the ***Supplementary Information*.** ECM components and architectural features were defined using National Centre for Biotechnology Information Medical Subject Heading terms. Non-cartilage soft joint tissues and disease nomenclature were also specified. The search strategy was validated against relevant papers identified in a preliminary literature search. The search strategy was run on the Ovid MEDLINE, Ovid EMBASE and Scopus platforms on 30 October 2020 and repeated on 1 October 2021.

### Eligibility criteria and screening

Abstracts were de-duplicated in Mendeley Reference Manager (Elsevier B.V., Netherlands) before importing into the Covidence platform. The remaining studies were screened independently at title/abstract and full-text stages by two reviewers (IR & JM), with conflicts resolves through consensus or a third reviewer (SS). Included studies were required to have ≥3 OA participants.

In human studies, eligible patients and controls were ≥18 years. Non-OA diseases, including inflammatory arthrides and crystalline arthropathies, were excluded. The presence of a valid control group was not a requirement for human studies. However, control groups were included if present and a minimum of 3 participants were included in this group. Valid control groups include tissues from healthy people or near-healthy tissues, including cadavers, individuals with osteosarcoma, and traumatic joint injuries provided that the comparator tissue was not directly damaged by the trauma.

In contrast to human studies, all animal studies required a control group. Studies that induced OA unilaterally and only used a contralateral control joint were excluded, as non-physiological loading of the contralateral joint induces ECM remodelling[21, 22]. Excluded animal models included the genetic deletion of ECM components, the introduction of matrix-degrading enzymes into the joint, surgical damage of a tissue subsequently reported on, and the ovariectomised rat model, as this is more commonly used as a model for osteoporosis[23, 24].

Regarding outcome measures, included studies evaluated at least one of the following tissues: intervertebral disc, ligament, skeletal muscle, tendon, meniscus, articular capsule, synovium, and fat pad. Papers that only studied these tissues after treatment, including – but not limited to – surgical or drug treatment, or after these tissues were purposely injured to induce the development of OA, were excluded. Papers evaluating non-ECM tissue components (cells, cytokines, matrix-degrading enzymes) were ineligible for inclusion. Given the focus on structural ECM, regulatory matricellular proteins, as well as neoepitopes generated during ECM turnover, were not included. Studies using *in vitro* or *ex vivo* culture systems were excluded as the ECM proteins cells synthesise differ in culture and *in vivo*. Transcriptomic analyses were excluded as gene expression is a determinant, not a measure, of protein abundance. Finally, only English-language articles were included.

### Data extraction and bias analysis

Data were extracted from all included studies by one reviewer (JM or IR) using a standardised extraction form in Microsoft Excel; the extraction was verified by the other reviewer. Where there was uncertainty, extraction was performed in duplicate by both reviewers. Number of participants (or animals) in each group was recorded as well as the presence/absence of a control group; if a control group was present, the control population and control tissue were described. For animal studies, the species, strain, and type of OA model were recorded. When available, participant age, sex, body mass index (BMI) and disease severity were recorded, as were the joint and tissue being studied. Relevant ECM components and architectural features were described; comparisons to control tissues and statistical analysis were noted when applicable. Results were grouped by tissue, followed by ECM feature, and finally the direction of change compared to control (increase, no change, decrease, or no control group present) and presented in Tables 1 (human studies) and 2 (animal studies). Due to the large number of different included ECM features, accepted research methods, and accepted measures of effect, a quantitative meta-analysis was not deemed appropriate. Bias analysis was performed by IR, with all included studies assessed using the 2015 Office of Health Assessment and Translation (OHAT) Risk of Bias Rating Tool for Human and Animal Studies. The results of the bias analysis can be found in Supplementary Table 3.

## 3 Results

### 3.1. Study overview

18,133 potentially relevant articles were identified by the search strategy (Figure 1). Following the removal of duplicates, 8,378 abstracts were screened. Of the 399 studies assessed for eligibility at full-text screening, 142 met all criteria for inclusion in this review. The characteristics of all included studies are summarised in ***Supplementary Tables 1 and 2 (human and animal studies, respectively).*** A schematic overview of the included studies can be found in Figure 2.

**Figure 1.**
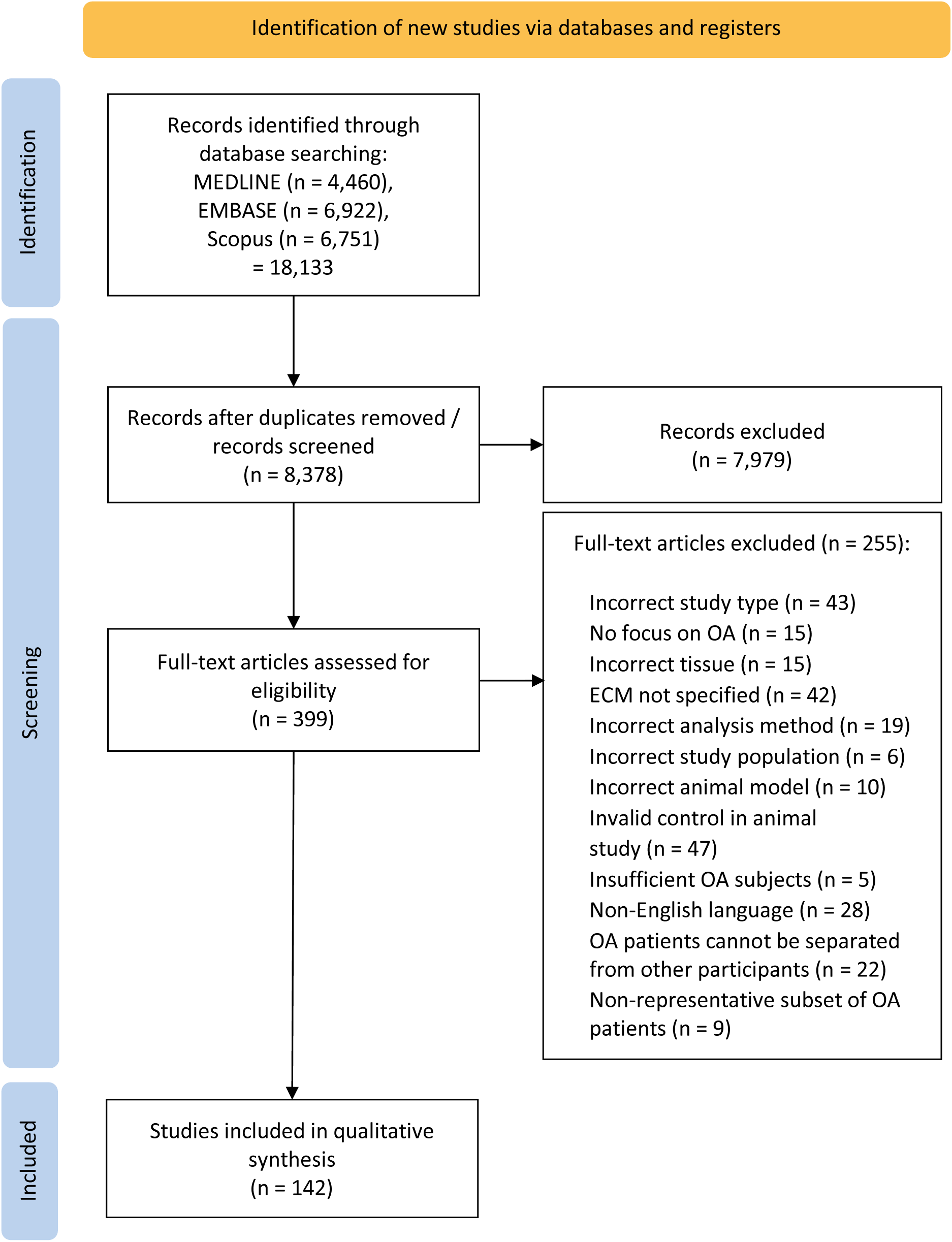
PRISMA 2022 flow diagram.

**Figure 2.**
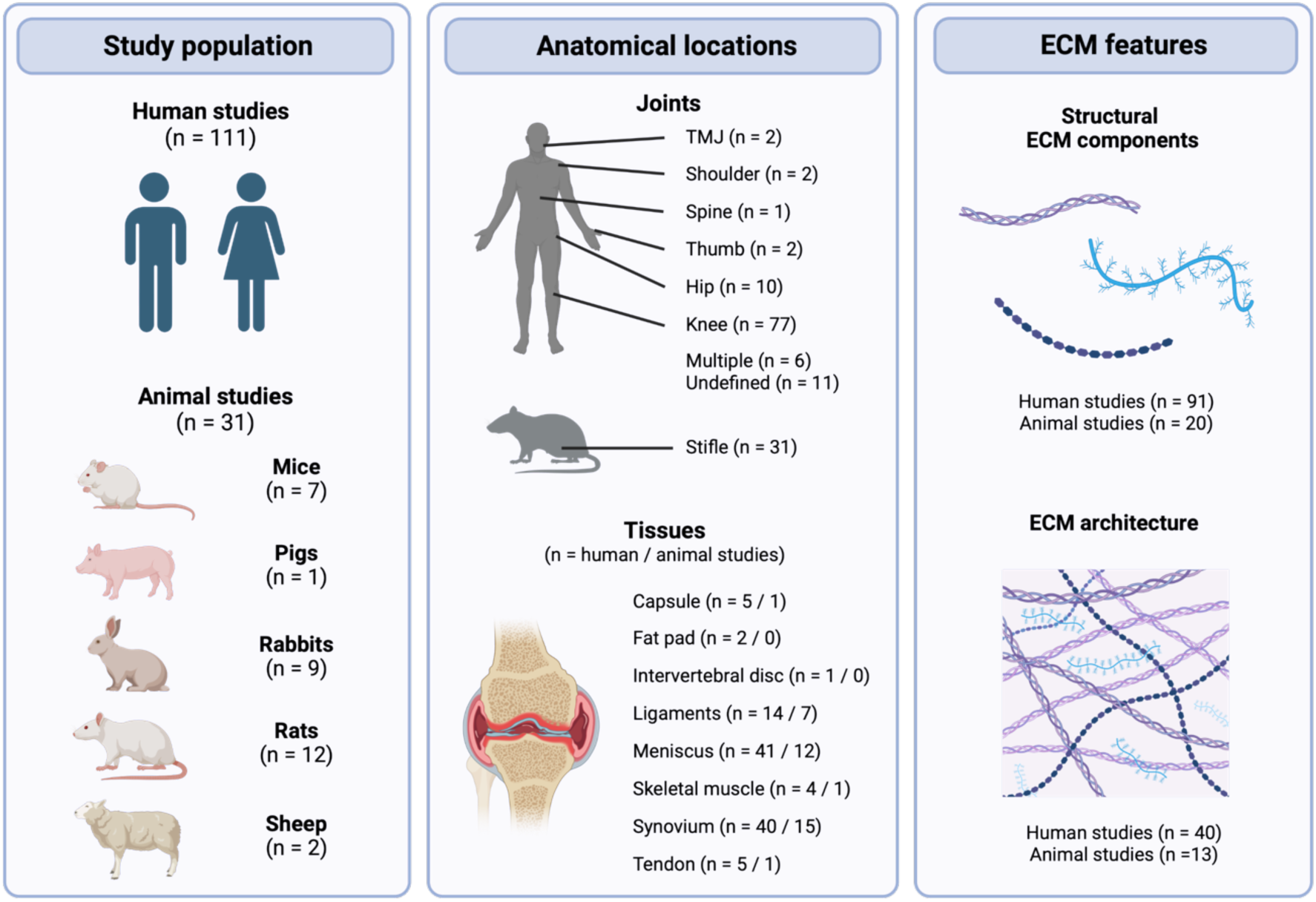
Schematic overview of the study population, anatomical locations, and ECM features studied in the included studies. Created with BioRender.com.

### 3.2 Human studies

Most studies investigated meniscus (n = 41) and synovium (n = 40), followed by ligaments (n = 14), tendon (n = 5), capsule (n = 5), skeletal muscle (n = 4), fat pad (n = 2) and intervertebral disc (n = 1) (Supplementary Table 1). Studies mostly commonly investigated the knee joint (n = 77), but papers on hip (n = 10), thumb (n = 2), temporomandibular joint (TMJ) (n = 2), shoulder (n = 2), and spine OA (n = 1) were also identified. While most studies on synovium, tendon, and capsule focussed on the presence/absence and distribution of specific ECM components, a large proportion of the papers on meniscus and ligaments investigated ECM architecture and viscoelastic properties (Table 1).

**Table 1.** Structural ECM components and architectural features in non-cartilage soft tissues of human osteoarthritic joints. **Abbreviations:** AA = amino acid, ACL = anterior cruciate ligament, CILP = cartilage intermediate layer protein, CHS = chondroitin sulphate, CPPD = calcium pyrophosphate deposition, CT = computed tomography, DS = dermatan sulphate, ECM = extracellular matrix, EDA = ethylenediamine, ED + F = enzymatic digestion and fractionation, EM = electron microscopy, FCCIS = fine cationic colloidal iron staining, FITR = Fourier-transformed infrared spectroscopy, Fn = fibronectin, F-UAA-EP = Fractination - uronic acid assay – electrophoresis, GAG = glycosaminoglycan, HA = hyaluronic acid, H&E = haematoxylin and eosin, HS = heparan sulphate, IB = immunoblotting, IEC = ion exchange chromatography, IHC = immunohistochemistry, IP = immunoprecipitation, IRT = indentation relaxation tests, KL-grade = Kellgren-Lawrence grade, LC = liquid chromatography, LM = light microscopy, N/A = not applicable, NI = nanoindentation, NR = not reported, OA = osteoarthritis, PAS = periodic acid Schiff, PCL = posterior cruciate ligament, PLM = polarised light microscopy, qual = qualitative, quant = quantitative, s-quant = semi-quantitative, SPA = sandwich-binding protein assay, TEM = transmission electron microscopy, WB = western blot.

| Tissue | ECM feature | Change vs control | Description of feature in OA tissue | Measurement technique(s) | Analysis | Stat. test | Ref |
| --- | --- | --- | --- | --- | --- | --- | --- |
| Capsule | Calcification | No control | Minute shards of calcific detritus commonly abutted on the degenerative areas. | NR | Qual | N/A | [27] |
|  | Collagen content | No control | Capsule tissue is mostly collagenous (range 15.6-88.8% of area; average 57.0% and 70.4% in two different OA groups) | Trichrome | Qual | N/A | [26] |
|  | Collagen cross-links | ↑ | All OA samples contained reducible cross-links not seen (or in minor quantities) in the joint capsule of healthy adults, namely intermediate Schiff-base cross-links. | IEC | Quant | No | [29] |
|  | Collagen fibre organisation | ↓ | Disordered compared to control: the normal, smooth, orderly arrangement of collagen fibres is lost and replaced by a jumbled mass like a tangled klein of wool. | EM | Qual | N/A | [25] |
|  | ECM organisation | No control | Matrix appeared pale and more fibrillary and whorled in degenerated than in non-degenerated chondroid areas. | EM, Safranin O-Fast Green | Qual | N/A | [27] |
|  | Elastic fibres | No control | Degenerated areas were more intensely stained than the non-degenerated chondroid areas. | Adehyde-fuchsin | Qual | N/A | [27] |
|  | GAG / proteoglycan | No control | Degenerated areas were less intensely stained with Safranin O | Safranin O | Qual | N/A | [27] |
|  | GAG: Chondroitin sulphate (CHS) | No control | Chondroitin-6-sulphate (CHS-C) present in joint capsule; major polysaccharide component together with DS and HA. Ratio of CHS-C to DS of approximately 1:1. | ED + F | Quant | N/A | [28] |
|  | GAG: Dermatan sulphate (DS) | No control | Present in joint capsule; major polysaccharide component together with CHS-C and HA. Ratio of CHS to DS of approximately 1:1. | ED + F | Quant | N/A | [28] |
|  | GAG: Hyaluronic acid (HA) | No control | Present in joint capsule. Major polysaccharide component together with CHS-C and DS. | ED + F | Quant | N/A | [28] |
|  | GAG: Hexosamine | No control | ~1.1% hexosamine concentration in joint capsule, of which ~80% was glucosamine and ~20% was galactosamine | ED + F | Quant | N/A | [28] |
| Fat pad | Collagen I | ↓ | From ~0.6% in controls to ~0.1% area stained in end-stage OA | Sirius red | Quant | Yes | [31] |
|  | Collagen III | ↓ | From ~1.6% in controls to ~0.4% area stained in end-stage OA | Sirius red | Quant | Yes | [31] |
|  | COMP | = | Variable staining, more intense in fibrous than adipose regions of fat pad | IHC, IB | S-quant | Yes | [30] |
| Intervertebral disc | Calcification | No control | Increasing calcification with increasing OA severity | CT | Quant | N/A | [32] |
| Ligament | Aggrecan | No control | Weak expression around cells in OA ACL and PCL, only in higher grades of degeneration | IHC | Qual | N/A | [37] |
|  | Calcification | ↑ | Present in 3-31% of ACL tissue area | not specified | Quant | No | [36] |
|  | Collagen content | = | 63.4% (femoral region); 65.1% (midportion); 59.2% (tibial side); reduced collagen occupancy in PCL with increasing degree of OA | TEM; LM | Quant | Yes | [34] |
|  | Collagen I | No control | Present throughout most of ECM of ACL and PCL; prevalence similar with different degrees of degeneration | IHC | Qual | N/A | [37] |
|  | Collagen II | ↑ | Present in areas of fibrocartilaginous metaplasia in ACL; appears increased | IF | Qual | N/A | [36] |
|  |  | No control | Focal deposits found around cells in OA ACL and PCL, only present in higher grades of degeneration | IHC | Qual | N/A | [37] |
|  | Collagen III | No control | Present in ECM surrounding cells in early and advanced degeneration in OA ACL and PCL | IHC | Qual | N/A | [37] |
|  | Collagen fibre density | No control | Mean fibre density in ACL: 142.51/μm<sup>2</sup> | TEM | Qual | N/A | [44] |
|  | Collagen fibre diameter | = | 84.0nm (femoral region); 81.6nm (midportion); 85.5nm (tibial region). Fibril diameter of PCL smaller in OA patients with marked degeneration (75.1 nm) than those with less marked degeneration (87.5 nm) | TEM | Quant | Yes | [34] |
|  |  | No control | Fibres most commonly 50-100nm in diameter (0-50nm: 9.0%, 50-100nm: 60.6%, 100-150nm: 19.5%, 150-200nm: 10.8%, >200nm: 0.0%) in ACL | SEM/TEM | Qual | N/A | [44] |
|  | Collagen fibre organisation | ↓ | Organisation in ACL and PCL decreases with increased OA severity; fibre disorganisation higher in ACL than PCL | H&E + LM | S-quant | Yes | [38] |
|  |  |  | Collagen fibre degeneration in PCL positively correlated with severity of OA | Van Gieson and Gomori | S-quant | Yes | [39] |
|  |  |  | Collagen fibres in ACL frequently disoriented compared to normal group | H&E + LM | Qual | N/A | [41] |
|  |  | No control | Disorganised (ACL & PCL) | H&E + LM | Qual | N/A | [33] |
|  |  |  | Range from normal to degeneration in >2/3 of tissue (ACL & PCL); ACL more likely to have significant degeneration (no stats test); positive correlation between ACL appearance and collagen degeneration in PCL | H&E + LM | S-quant | N/A | [35] |
|  |  |  | Increasingly severe OA associated with reduced collagen organisation at bony insertion of palmar beak ligament | H&E + LM | Qual | N/A | [45] |
|  |  |  | Highly variable in ACL and PCL; reduced organisation seen as grade of ligament degeneration increased | H&E + LM | Qual | N/A | [37] |
|  |  |  | Most PCLs (65.2%) showed severe collagen fibre degeneration; no correlation between OA grade and fibre degeneration | Gomori trichrome & H&E | S-quant | N/A | [40] |
|  |  |  | Dorsoradial ligament: organized collagen bundles with collinear orientation. Anterior oblique ligament: disorganized connective tissue with few collagen fibers, appearance similar to synovial tissue | IF | Qual | N/A | [46] |
|  |  |  | Range from parallel to irregular in PCL; organisation decreased with higher OA grades | von Gieson stain + LM | S-quant | N/A | [42] |
|  |  |  | Organisation normal in 79% of PCL samples | H&E/Alcian blue + LM | S-quant | N/A | [43] |
|  |  |  | Collagen fibres in ACL largely parallel but disordered particularly at bony insertions | Toluidene blue + H&E | Qual | N/A | [44] |
|  | Decorin | No control | Greatest staining in the middle region of the ACL | IHC | Qual | N/A | [44] |
|  | GAG / proteoglycan | ↑ | Increased in ACL, especially in areas of cartilaginous metaplasia | Safranin-O | Qual | N/A | [36] |
|  |  | No control | Present around cells in ACL and PCL | Alcian blue | Qual | N/A | [37] |
| Meniscus | Aggrecan | ↑ | Strong (significant) increase in staining, which was more prominent in the deep zone than the surface zone | IHC | S-quant | Yes | [83] |
|  |  | No control | Highest density of staining around cell clusters, followed by staining in degenerative areas, and finally staining in intact areas. | IHC | Quant | N/A | [65] |
|  | Biglycan | = | No change in fragmentation compared to age-matched controls | WB | Qual | N/A | [70] |
|  | Calcification | ↑ | Extensive calcified areas; increase in size and frequency compared to control | Alizarin red | Qual | N/A | [49] |
|  |  |  | Significant increase in calcification in both medial (11.9% vs 5.2%) and lateral regions (11.1% vs 4.2%), as well as both menisci (11.5% vs 4.7% of surface area) | H&E | Quant | Yes | [69] |
|  |  | ↑/= | Increase in lateral anterior and medial anterior regions. Non-significant increase in lateral posterior and non-significant decrease in medial posterior regions. | Von Kossa | S-quant | Yes | [47] |
|  |  | No control | Heterogeneous distribution, indicating micro-calcifications; contained within both apatite and calcium pyrophosphate crystals in cartilaginous areas | X-ray AS, X-ray F, FTIS | Quant | N/A | [50] |
|  |  |  | Calcified areas found | Alizarin red | Qual | N/A | [60] |
|  |  |  | Presence of mineral deposition; approximately 80% of mineral areas co-localised with CPPD crystals. | Alizarin Red + Eosin Y | Qual | N/A | [64] |
|  |  |  | Calcium deposits found in 16% of OA menisci | IHC | Qual | N/A | [67] |
|  |  |  | Increase in calcification in menisci with complete tears vs menisci with no or partial tears | Alizarin Red | Quant | N/A | [75] |
|  |  |  | Calcification in all OA menisci, but not the two adults in normal group. Small/medium-sized deposits at the edges of sections (almost all patients); clusters of small-sized deposits inside the meniscus (65% of patients); and widespread medium/large-sized deposits (35% of patients). | Alizarin red | S-quant | Yes | [82] |
|  |  |  | 4/18 patients had calcification | X-ray | Qual | N/A | [84] |
|  |  |  | Extensive meniscal calcification | H&E | Qual | N/A | [87] |
|  | Cartilage intermediate layer protein (CILP) | ↑ | CILP was abundant in the matrix separating lacunae of the central region, but undetectable in peripheral regions. Sparse expression in areas of apoptosis and calcification. | IHC | Qual | N/A | [60] |
|  | Collagen content | ↑ | Significant increase in collagen content compared to control | Colorimetry | Quant | Yes | [78] |
|  |  | = | No difference in hydroxyproline content (major component of collagen) | Amino acid analyser system | Quant | Yes | [84] |
|  |  | ↓ | Statistically significant decrease in staining in OA (grade 0-2) compared to normal (grade 2) | Picrosirius red | S-quant | Yes | [83] |
|  |  |  | Significant decrease in collagen content in degenerated vs non-degenerated areas | Colorimetry | Quant | N/A | [54] |
|  |  | No control | Collagen per dry mass did not show much variation among anterior, central, and posterior regions but varied in the radial direction: outer region was higher than other regions. Lower content in medial than lateral meniscus. Collagen per wet mass showed little variation. | Spectrophotometry | Quant | N/A | [81] |
|  |  |  | Collagen content decreased from no to high degeneration; no differences between regions. | IR spectroscopy | S-quant | N/A | [86] |
|  | Collagen cross-links | ↑/↓ | Significant decrease in mature cross-links pyridinoline and deoxypyridinoline in OA compared to controls. Pentosidine (senescent cross-link) was non-significantly increased. | HPLC | Quant | Yes | [84] |
|  | Collagen I | ↓ | Significant decreased throughout meniscus, particularly in the middle zone and deep zones, indicating greater decrease in the middle and deep zones than in the surface zones. | IHC | S-quant | Yes | [83] |
|  |  | No control | Non-degenerated menisci: expression varied by subject; expression throughout tissue except peripheral vascular area; intense expression in circumferential and radial fibres. Degenerated menisci: expression decreased in degenerated areas and was almost absent in severely degenerated menisci. | IHC | Quant | N/A | [71] |
|  |  |  | Decrease in staining with increase in damage (no tear > partial tear > complete tear). | IHC | Quant | N/A | [75] |
|  |  |  | Expression decreased in menisci that showed degeneration (compared to samples without degeneration), particularly in the body of meniscus using a S-quantitative score, but not using a quantitative score. | IHC | S-quant / quant | N/A | [80] |
|  | Collagen II | ↓ | Significant prominent decrease in type II collagen in surface, middle, deep zones. | IHC | S-quant | Yes | [83] |
|  |  | No control | Highest to lowest expression: anterior, central, and posterior region. | IHC | Quant | N/A | [56] |
|  |  |  | Highest density of staining around cell clusters, followed by staining in degenerative areas, and finally staining in intact areas. | IHC | Quant | N/A | [65] |
|  |  |  | Non-degenerated menisci: expression similar between subjects; intense expression at exterior peripheral border, on meniscal surface, and along vessels but not in any internal region that expressed collagen II. Degenerated menisci: expression decreased in degenerated areas but increased slightly in degenerative menisci; expression almost absent in severely degenerative menisci. | IHC | Quant | N/A | [71] |
|  |  |  | Increase in staining with increase in damage (no tear > partial tear > complete tear). | IHC | Quant | N/A | [75] |
|  | Collagen III | No control | Non-degenerated menisci: expression similar between subjects; expression throughout tissue except peripheral vascular area; intense expression in circumferential and radial fibres. Degenerated menisci: expression decreased in degenerated areas and was almost absent in severely degenerative menisci. | IHC | Quant | N/A | [71] |
|  | Collagen X | No control | Positive staining in superficial layers | IHC | Qual | N/A | [57] |
|  |  |  | High levels throughout the tissue; co-localised with calcium mineral staining | IHC | Qual | N/A | [64] |
|  | Collagen fibre diameter | ↓/= | Collagen fibril diameter was decreased and number of fibrils per area was significantly increased in medial menisci, but not in lateral menisci. | TEM | Quant | Yes | [63] |
|  |  | ↓ | Decreased diameter in OA compared to control: 35-45 nm in OA compared to 70-80 nm in controls | TEM | Quant | No | [49] |
|  |  | No control | Average transverse collagen diameter was 95.39 +- 15.87 nm and longitudinal diameter was 96.3 +- 14 nm for control method | TEM | Quant | N/A | [58] |
|  | Collagen fibre organisation | ↓/= | Percentage of area occupied by fibrils was reduced in medial but not in lateral menisci. | TEM | Quant | Yes | [63] |
|  |  | ↓ | Disorganized collagen fibres in OA; collagen fibres were aligned with a regular pattern of distribution and diameter in controls | TEM | S-quant | No | [49] |
|  |  |  | Organised collagen network was lost in medial OA meniscus; medial OA meniscus 24-25 degrees lower orientation angles compared to lateral OA or donor menisci | μCT | Quant | N/A | [61] |
|  |  |  | Lateral menisci: fibril bundles tended to be obscure in the surface regions, but were well preserved in the inner area. Medial menisci: less even distribution of fibril bundles; tended to be thicker than controls and were not clearly recognised in the surface regions; obvious wearing and fraying in degenerated areas. | H&E, Masson's trichrome, | Qual | N/A | [63] |
|  |  |  | Some separation of individual collagen fibre bundles in patients without a tear; disorganised collagen architecture in patients with a partial tear; collagen architecture completely disorganised. | Safranin O, H&E, Picrosirius red | Qual | N/A | [75] |
|  |  |  | Collagen networks were less organised and less compact than the well-organised, parallel-arranged, and compact collagen bundles in normal menisci. | Picrosirius red, H&E | Qual | N/A | [83] |
|  |  | No control | Collagen fibres were separated and concentrated in the OA meniscus. Also homogenised collagen fibres surrounding cells | H&E | Qual | N/A | [48] |
|  |  |  | Fibre angle compared to tidemark remained constant in medial entheses but increase in lateral entheses as tissue transitioned from ligament zone to subchondral bone. Significant differences in fibre angle and deviation in calcified fibrocartilage of lateral meniscus. | Picrosirius red + PLM | Quant | N/A | [55] |
|  |  |  | Tears in collagen fibres found in all studied OA subjects | H&E | Qual | N/A | [74] |
|  |  |  | A collection of transverse collagen fibres (transverse ligament) were found running from the synovial edge into the vascular region, which presented a "tree-like" formation of fibres interwoven into the avascular region in all samples | H&E | Qual | N/A | [85] |
|  | COMP | No control | Significant decrease in staining: in OA tissue the median staining of ECM was 0 (range 0-1); it was 1 (range 0.66-3) in tissue from meniscal horn tears | IHC | S-quant | Yes | [67] |
|  | Compressive modulus | ↓ | Instantaneous and equilibrium elastic modulus decreased, particularly in lateral anterior and medial anterior regions | NI | Quant | Yes | [47] |
|  |  |  | Aggregate modulus of OA medial menisci was 40% lower than that of control medial menisci, whereas it differed little in lateral menisci | Indentation | Quant | Yes | [63] |
|  |  | No control | Lateral anterior, lateral posterior, and medial anterior regions showed decreasing instantaneous and equilibrium moduli with increasing degeneration. The medial posterior region had no apparent trend in mechanical properties with degeneration | IRT | Quant | N/A | [51] |
|  | Decorin | ↑ | Increase in fragmentation compared to age-matched controls | WB | Qual | N/A | [70] |
|  | Fibromodulin | = | No change in fragmentation compared to age-matched controls | WB | Qual | N/A | [70] |
|  | GAG /<br>proteoglycan | ↑ | Increase in GAG compared to control | Safranin O/fast green | Qual | N/A | [49] |
|  |  |  | Intense proteoglycan staining in OA, weak staining in healthy tissue | Safranin O/Fast Green | Qual | N/A | [66] |
|  |  |  | Increase in median Safranin-O score throughout the whole meniscus (3 vs 1 in control). | Safranin O | S-quant | Yes | [77] |
|  |  |  | Significant increase in proteoglycan content (overall, medial, and lateral) compared to controls; significant increase in medial compared to lateral OA menisci. | Spectrophotometric analysis | Quant | Yes | [78] |
|  |  |  | Significantly stronger proteoglycan staining in OA than normal menisci. The increase of safranin-O staining was more prominent in the deep zone than that in the surface zone | Safranin-O Fast green, Alcian Blue, Toluidine Blue | S-quant | Yes | [83] |
|  |  | No control | Zone 1 (basis to the most peripheral blood vessel): mostly neutral carbohydrates but acid mucopolysaccharide around vessels. Zones 2 & 3 (peripheral blood vessel to the centre in two halves): zone 2 mostly acid mucopolysaccharide; zone 3 mix of neutral carbohydrates and acid mucopolysaccharides, with acid mucopolysaccharides in central meniscus, neutral carbohydrates at the top and some edges. | Alcian blue/PAS | S-quant | N/A | [53] |
|  |  |  | Higher proteoglycan staining in anterior 1/3 than central or posterior thirds | Safranin O | Quant | N/A | [56] |
|  |  |  | Weak proteoglycan staining in superficial regions | Safranin O | Qual | N/A | [57] |
|  |  |  | Specific proteoglycan staining in/around meniscal cell clusters | Safranin O | Qual | N/A | [65] |
|  |  |  | 42% of OA menisci had none/weak staining. | Alcian blue | S-quant | No | [67] |
|  |  |  | Non-degenerated: GAG expression on surface of meniscus, vessels, and internal regions where collagen II was expressed. Degenerated: increase in GAG expression. | IHC | Quant | N/A | [71] |
| | | | Proteoglycan score $2.7 \pm 0.4$ (score 0-3) | Safranin O-Fast Green | S-quant | N/A | [72] |
|  |  |  | Little variation in sGAG per dry mass among anterior, central, and posterior regions. Outer and middle regions were higher than inner region. Higher in medial than lateral meniscus. sGAG per wet mass was higher in medial meniscus for the anterior and central regions, but not the posterior region. Content highest to lowest: middle > outer > inner region. Staining confirmed the greatest intensity in middle and lowest inner regions. | DMMB assay / Safranin-O | Quant | N/A | [81] |
|  |  |  | Significantly more GAG staining in vascular than avascular areas of meniscus. The inner borders stained more strongly than the other zones. | Toluidine blue | S-quant | N/A | [85] |
|  |  |  | Non-significant decrease in proteoglycan content with increasing degeneration; no differences between regions. | IR spectroscopy | Quant | N/A | [86] |
|  | GAG thickness | ↑/= | Increased in the lateral anterior, medial anterior and medial posterior regions of calcified zones; no difference in uncalcified zones | Toluidine blue | Quant | Yes | [47] |
|  | GAG: chondroitin sulphate (CHS) | = | CHS-A and CHS-C present. OA menisci showed same composition as those from a comparable age group. | F-UAA-EP | Quant | No | [62] |
|  | GAG: dermatan sulphate (DS) | = | Present. OA menisci showed same composition as those from a comparable age group. | F-UAA-EP | Quant | No | [62] |
|  |  | No control | Staining present in fibrous matrix of the superficial layer | IHC | Qual | N/A | [68] |
|  | GAG: Hexosamines | No control | Significant increase in hexosamine content in degenerated vs non-degenerated areas | Colorimetry | Quant | N/A | [54] |
|  | GAG: Hyaluronic acid (HA) | = | Present. OA menisci showed same composition as those from a comparable age group. | F-UAA-EP | Quant | No | [62] |
|  |  | No control | Strong staining in vascular zone, particularly in intima and subintima, in the vicinity of blood vessels and fatty tissue; moderate/weak staining in the inner zones | IHC | S-quant | N/A | [53] |
|  | GAG: heparan sulphate (HS) | = | Present. OA menisci showed same composition as those from a comparable age group. | F-UAA-EP | Quant | No | [62] |
|  | GAG: Keratan sulphate (KS) | = | Present. OA menisci showed same composition as those from a comparable age group. | F-UAA-EP | Quant | No | [62] |
|  | Hydroxyproline | = | No difference in hydroxyproline content | AA analyser system | Quant | Yes | [84] |
|  | Keratocan | = | No change in fragmentation compared to age-matched controls | WB | Qual | N/A | [70] |
|  | Lubricin | ↓ | Weak expression in OA (moderate expression in outer and middle rim, and weak expression in inner rim), while expression was strong in normal donors in all compartments. | IHC | S-quant | No | [73] |
|  |  | No control | Staining in all patients, ranging from weak to very strong. Matrix staining strongest directly beneath the surface and decreasing in intensity in deeper tissue. Discrete granular deposits observed. Staining followed crimp and trabecular patterns of collagen fibrils. | IHC | S-quant | N/A | [87] |
|  | Lumican | = | No change in fragmentation compared to age-matched controls | WB | Qual | N/A | [70] |
| | Viscoelastic properties | ↑ | Wide distribution of elastic moduli demonstrates stiffer mean values in the outer ($364.3 \pm 72.2$ kPa), middle ($401.9 \pm 39.2$ kPa), and inner regions ($365.9 \pm 106.5$), compared to $303.0 \pm 28.3$ kPa, $282.4 \pm 30.2$ kPa, and $274 \pm 8.7$ kPa, respectively, in aged normal donors | AFM | Quant | No | [66] |
| | | ↑/= | Non-significant ($p = 0.06$) increase in instantaneous modulus in posterior horn ($0.17 \pm 0.1$ MPa vs $0.12 \pm 0.07$ MPa in control) and entire meniscus ($0.17 \pm 0.07$ MPa vs $0.14 \pm 0.08$ MPa in control), but significant increase in maximum applied load for both. | Indentation test | Quant | Yes | [77] |
|  |  | =/↓ | Aggregate modulus of OA medial menisci was 40% lower than that of control medial menisci, whereas it differed little in lateral menisci | Indentation | Quant | Yes | [63] |
|  |  | ↓ | Instantaneous and equilibrium elastic modulus decreased, particularly in lateral anterior and medial anterior regions | NI | Quant | Yes | [47] |
|  |  | No control | Lateral anterior, lateral posterior, and medial anterior regions showed decreasing instantaneous and equilibrium moduli with increasing degeneration. The medial posterior region had no apparent trend in mechanical properties with degeneration | IRT | Quant | N/A | [51] |
|  |  |  | No significant differences were found between degenerative groups for the tensile modulus. Average tensile modulus: lateral anterior 128.8±62.5 MPa, lateral posterior 119.4±75.6 MPa, medial anterior 112.5±56.6 MPa, and medial posterior 95.9±55.6 MPa. Regardless of degenerative grade or region the average strain at failure was 18.2±5.2%. | Pulling to failure | Quant | N/A | [51] |
|  |  |  | Tensile elasticity modulus was 11.66 MPa for control method (cryoprotective preservation). The stress-strain curve revealed a slow-slope curve with a non-abrupt rupture (ductile material). Different results were found for other methods of preservation | Tensile testing machine | Quant | N/A | [59] |
|  |  |  | The average values for equilibrium compressive modulus, dynamic compressive modulus, and dynamic shear modulus were 52.2 (39.9, 72.6) kPa, 212.3 (157.8, 282.3) kPa, and 15.5 (12.0, 20.1) kPa, shown as mean (95% upper limit, 95% lower limit) respectively. Overall, mechanical properties exhibited little regional variation. | Torsional rheometer / Microtester | Quant | N/A | [81] |
|  |  |  | Equilibrium modulus decreases with increased degeneration (not dependent on region). Hydraulic permeability increased 50% from no to high degeneration, with the anterior being lower than intermedia and posterior regions. Non-significant decrease in tensile modulus from no to high degeneration; no difference between regions. | Compression / tensile test | Quant | N/A | [86] |
|  | ECM composition (proteomics) | ↑/=/↓ | Differences in medial region, but not in lateral region. Full details in paper. | MS | Quant | Yes | [52] |
|  |  |  | Both in- and decreases found in ECM components in OA compared to control. Full details in paper. | MS | Quant | No | [79] |
|  |  | No control | 131 ECM or ECM-associated proteins were identified in meniscal tissue by MatrisomeDB 2.0. COMP, CILP, aggrecan, and 7 collagens were dominantly identified in both lateral and medial menisci. 14 ECM proteins were differentially expressed between medial and lateral meniscus. Full details in paper. | MS | Quant | N/A | [76] |
| Skeletal muscle | Calcification | No control | Marked calcification of perimysium & juxtavascular tissue of the vastus medialis muscle | H&E + LM | Qual | N/A | [88] |
|  | Collagen content | ↑ | Collagen content higher in OA vastus lateralis muscle vs controls and negatively correlated with muscle strength | Sirius red | Quant | Yes | [90] |
|  | Collagen I | ↑ | Weak to moderate staining of OA vastus lateralis muscles, appears increased compared to control | IHC | S-quant | No | [91] |
|  |  | No control | All vastus lateralis muscles stained at least weakly positive | IHC | S-quant | N/A | [89] |
|  | Collagen III | ↑ | Light to strong staining of OA vastus lateralis muscles, appears increased compared to control | IHC | S-quant | No | [91] |
|  |  | No control | All vastus lateralis muscles stained at least weakly positive | IHC | S-quant | N/A | [89] |
|  | Collagen IV | ↑ | Weak to strong staining in OA vastus lateralis muscles, appears increased compared to control | IHC | S-quant | No | [91] |
|  |  | No control | All vastus lateralis muscles stained at least weakly positive | IHC | S-quant | N/A | [89] |
|  | GAG / proteoglycan | ↑ | Increased GAG content in OA vastus lateralis muscle vs control; GAG co-localised with collagen | Wheat germ agglutinin | Quant | Yes | [90] |
|  | Laminin | No control | Present in the basement membrane of OA vastus lateralis muscles | IHC | Qual | N/A | [91] |
| Synovium | Calcification | ↑ | Presence of ectopic calcification in OA, but not control samples | H&E | Qual | N/A | [103] |
|  |  | No control | Calcium deposits found in synovium of nearly all KL-grades, including KL 0 | Alicarin red, FITR | Qual | N/A | [95] |
|  |  |  | Primary OA: calcification present in subsynovial foci in cases with minimal hyperplasia Rapidly destructive OA: ossification was well defined in the sublining<br>Secondary OA: large areas of calcification were observed. | H&E | Qual | N/A | [112] |
|  |  |  | Calcium was found in most OA patients (14/16), mostly located close to the synovial surface; ~1% of the relative volume of the synovium. | IHC (Alizarin red) | S-quant | N/A | [117] |
|  | Collagen content | No control | Found in all OA patients; ~15.37% of the volume of synovium | Movat's pentachrome | S-quant | N/A | [117] |
|  | Collagen I | ↑ | Expression present and increased in sublining; present but unchanged in lining | IF | Qual | N/A | [128] |
|  |  | No control | Expression in synoviocytes, their matrix, and the intimal layer of small vessels of the synovium. | IHC | Qual | N/A | [68] |
|  | Collagen II | No control | Small cartilage particles found in OA synovium | IHC | Qual | N/A | [124] |
|  |  |  | Collagen 2-positive fragments surrounded by lining cells of villi in all OA synovial specimens; on average 9.1 fragments (range 3-26) in each x400 magnification field | IHC | S-quant | N/A | [100] |
|  |  |  | Cartilaginous fragments of varying sizes entrapped between synovial lining cells. | IHC | S-quant | N/A | [105] |
|  |  |  | Cartilage/bone fragments were found in 3 out of 16 samples. | unclear | Qual | N/A | [117] |
|  | Collagen III | ↑ | Present in sublining | IF | Qual | N/A | [128] |
|  |  | No control | Intense nitrated type III collagen expression found in the basement membranes of blood vessels and the lining layer within the fibrous tissue surrounding synoviocytes | IHC | Qual | N/A | [104] |
|  |  |  | Expression found in vascular walls and in surrounding connective tissue matrix | IF | Qual | N/A | [129] |
|  | Collagen IV | ↑ | Present in sublining | IF | Qual | N/A | [128] |
|  |  | = | Present in basement membranes of vasculature and pericellular distribution in the intimal layer but not underlying connective tissue. No change compared to trauma control. | IHC | Qual | N/A | [114] |
|  |  | No control | Present in sublining | IHC | Qual | N/A | [110] |
|  |  |  | Present in basement membrane of blood vessels | IF | Qual | N/A | [129] |
|  | Collagen V | ↑ | Present in sublining | IF | Qual | N/A | [128] |
|  | Collagen fibre architecture | No control | Accumulation of collagen fibres in early OA; collagen fibres become disordered in late OA. | H&E | Qual | N/A | [96] |
|  |  |  | Dense collagen bundles present in sublining; increasingly so with more severe OA. Giant collagen fibrils occasionally found in more severely degraded sublining. | EM, LM | Qual | N/A | [119] |
|  | Collagen cross-links | No control | Pyridinoline (Pyr) and deoxypyridinoline (Dpy) ratio was 14.5+-6.2: Pyr 0.84 mol/mol collagen and Dpy 0.07 mol/mol collagen | LC | Quant | N/A | [98] |
|  |  |  | Pyr:Dpr ratio was 25.6 | LC | Quant | N/A | [106] |
|  | COMP | No control | Expressed in and around synovial cells, less expression in sublining | IHC | Qual | N/A | [94] |
|  | Elastin | No control | Intense staining in the internal elastic lamina | IHC | Qual | N/A | [108] |
|  |  |  | Elastic fibrils increased in intermediate degeneration | EM, LM | Qual | N/A | [119] |
|  | Fibromodulin | = | Little fibromodulin expression found in OA synovial tissue. Tenancy towards lower expression than control, but not statistically significant | IHC | S-quant | Yes | [120] |
|  | Fibronectin (Fn)<br>(including isoforms) | ↑ | High expression areas close to blood vessels or high inflammation | MS & IHC | Quant | No | [92] |
|  |  |  | ED-A and ED-B-Fn mainly expressed in the lining; t-FN found throughout the lining and sublining | IHC | Qual | N/A | [93] |
|  |  |  | Fn expression in sublining | IF | Qual | N/A | [128] |
|  |  | No control | Small amount of Fn staining detected in sublining layer / Fn detected by WB | IHC & IP + WB | Quant | N/A | [121] |
|  |  |  | Little Fn staining found in deep sublining | IF | Qual | N/A | [97] |
|  |  |  | Weak staining of EDA+ Fn only in proliferative region of OA synovium | IHC | Qual | N/A | [122] |
|  |  |  | EDA+ Fn found in OA synovium; most intense staining in the lining layer | IF | Qual | N/A | [99] |
|  |  |  | Fn detected in lining, sublining, and blood vessels | IHC | Qual | N/A | [110] |
|  |  |  | tFn detected in lining, sublining, and blood vessels; EDA-Fn mainly in lining and vessels, limited expression in the sublining; expression of glycosylated-Fn was similar to EDA-Fn but with reduced intensity; EDB-Fn reduced expression in lining, few vessels stained, limited expression in sublining. | IHC | S-quant | N/A | [125] |
|  |  |  | Expression of Fn and EDA-Fn was strong in lining and blood vessels, but weak and irregular in the sublining | IHC | Qual | N/A | [111] |
|  |  |  | Fn expression found in lining layer, blood vessels, and area of high collagen content. | IHC | Qual | N/A | [113] |
|  |  |  | Fn expression found in lining layer | IHC | Qual | N/A | [101] |
|  |  |  | Expression of Fn in blood vessels and surrounding matrix. | IF | Qual | N/A | [129] |
|  |  |  | Patchy expression on synovial villi and in irregular ECM masses. | IF | Qual | N/A | [116] |
|  | GAG: Chondroitin sulphate (CHS) | ↑ | CHS-A/C expression increased in interstitial space below the lining layer compared to control. Present in perivascular and highly fibrous regions. | IHC (FCCIS) | Qual | N/A | [102] |
|  |  | ↓ | CHS-C absent from the lining layer in OA, while expression was found in normal synovium. Expression found in patches of faint staining in sublining, in a narrow band underlying vascular endothelium, and as a diffuse band outlining smooth muscle cells of the tunica media in larger vessels. | IHC | Qual | N/A | [118] |
|  | GAG: Dermatan sulphate (DS) | ↑ | Expression increased in interstitial space below the lining layer compared to control. Present in perivascular regions (in and around vascular endothelial cells) and subsurface interstitial space. | IHC (FCCIS) | Qual | N/A | [102] |
|  |  | = | Distributed through superficial and deep matrix; faint condensation in the adventitia. No differences with normal synovium reported. | IHC | Qual | N/A | [118] |
|  |  | No control | Expression found in synoviocytes and their surrounding matrix at the adventitial layer of small blood vessels | IHC | Qual | N/A | [68] |
|  | GAG: Hyaluronic acid (HA) | ↑ | HA found in blood vessels (at times extending into surrounding matrix), in areas of infiltrating cells, and reticular staining in the lining, which decreased upon hyperplasia. Low-to-moderate staining in OA compared to faint staining in normal tissue. | IHC | Qual | N/A | [130] |
|  |  | = | HA mainly found in lining, content varied strongly between patients; mean content (249 +- 34.8 µg/g) not significantly different from control (227.7 +- 35.4 µg/g). | IHC + SPA | Quant | Yes | [123] |
|  |  | No control | Staining was weak and restricted to the lining layer. | IHC | S-quant | N/A | [110] |
|  |  |  | Present in perivascular regions. | IHC (FCCIS) | Qual | N/A | [102] |
|  | GAG: Heparin sulphate (HS) | ↑ | HS expression increased in interstitial space below the lining layer compared to control. Present on and around vascular endothelial cells. | IHC (FCCIS) | Qual | N/A | [102] |
|  |  | No control | HS was detected with a diffuse pattern involving all vessels in OA sections | IF | Qual | N/A | [127] |
|  | Laminins | ↑ | EHS laminin (laminin-111) strongest expression in lining, areas of the sublining, and blood vessel basement membranes; occasional weaker staining in stromal pericellular areas. Moderate to strong staining for α5 and β1 (mainly in blood vessels but also in lining and sublining) and β2 laminin, which stained more extensively but less so in blood vessels. | IHC | S-quant | No | [109] |
|  |  | = | α2 and α3 laminin showed no/weak staining. No changed compared to trauma control. | IHC | S-quant | No | [109] |
| | | | α4 laminin showed weak expression in lining, strong staining in vascular basement membranes. | IF | Qual | N/A | [126] |
|  |  |  | EHS-laminin expression in basement membrane of vasculature and pericellular in intimal layer. | IHC | Qual | N/A | [114] |
|  |  |  | Laminin (unspecified) present in vascular basement membranes and areas of sublining with early connective tissue proliferation but not in mature fibrous tissue. | IF | Qual | N/A | [129] |
| | | ↓ | Weak α5 laminin expression in lining, strong staining in vascular basement membranes. | IF | Qual | N/A | [126] |
|  |  | No control | Pericellular distribution in the synovial lining layer. Expression appears to decrease in cases of severe inflammation | IHC | S-quant | N/A | [115] |
| | Latent TGF-β-binding protein 1 (LTBP-1) | ↑ | Increased expression (by IHC); 2-fold increase in expression (by WB). | IHC, WB | Quant | Yes | [107] |
|  | Lumican | = | High levels throughout synovial and subsynovial tissue | IHC | S-quant | NR | [120] |
|  | Reticulin | No control | Expression found in vascular walls and in surrounding connective tissue matrix | IF | Qual | N/A | [129] |
|  | Vitronectin | No control | Expression found in the lining layer and some expression is present in the sublining often in fibril like structures. | IHC | Qual | N/A | [101] |
| Tendon | Calcification | ↑ | Increase in internal obturator tendon calcification in OA compared to control | Von Kossa | S-quant | Yes | [135] |
|  |  | = / ↓ | Lower percentage of calcification in OA biceps but no difference in subscapularis tendon | Von Kossa + LM | S-quant | Yes | [132] |
|  |  | No control | Mean calcification score in OA Achilles tendon 2.79 (scored 0-6 for both tendons: 0=none, 1 = <5mm, 2 = 5-10mm, 3 = ossification >10mm) | Ultrasound | S-quant | N/A | [131] |
|  | Collagen I | = | Present in all OA long head of biceps tendons | IHC | S-quant | Yes | [134] |
|  | Collagen III | = | Present in all OA long head of biceps tendons | IHC | S-quant | Yes | [134] |
|  | Collagen fibre diameter | ↑ | Relatively fewer small and medium sized fibres (<30 & 31-60 nm) in OA internal obturator tendon vs control | TEM | Quant | Yes | [135] |
|  |  | ↑ / = | Increase in biceps (66.1nm vs 59.0nm in control); no changes in subscapularis (55.3nm vs 54.0nm in control) | H&E + LM, TEM | Quant | Yes | [132] |
|  |  | = | Mean diameter 68.3nm for in OA gluteus medius tendon (vs 67.6nm in control) | TEM | Quant | Yes | [133] |
|  | Collagen fibre organisation | = | Distal portion of long head of biceps tendon has more organised collagen than proximal; no difference with control | Polarised light | Quant | Yes | [134] |
|  |  | ↓ | Irregular collagen structure more common in OA biceps and subscapularis tendon vs control | TEM | S-quant | Yes | [132] |
|  |  |  | Slightly higher disorganisation of fibre structure in OA gluteus medius tendon than control | H&E + Alcian blue/PAS | S-quant | No | [133] |
|  |  |  | Irregular patterns (in ultrastructure) of collagen fibrils in OA internal obturator tendon compared to control | H&E + LM, TEM | Quant | Yes | [135] |
|  | Decorin | = | Present in all OA long head of biceps tendons | IHC | S-quant | Yes | [134] |
|  | GAG /<br>proteoglycan | ↑ | Greater proteoglycan content in OA long head of biceps tendon than control; greater proteoglycan content in proximal compared to distal tendon | Alcian blue | Quant | Yes | [134] |
|  |  |  | Increase in GAG between collagen fibrils in OA internal obturator tendon | Alcian blue | Quant | Yes | [135] |
|  |  | = | No difference in GAG in OA biceps and subscapularis tendon compared to control | Alcian blue | S-quant | Yes | [132] |
|  |  | ↓ | Slight decrease in average GAG score in OA gluteus medius tendon compared to control | Alcian blue/PAS | S-quant | No | [133] |

#### 3.2.1 Capsule in human OA

Of 5 studies which assessed the capsule (hip (n = 3), knee (n = 2)) [25–29], 4 were published before the year 2000. These studies covered both ECM components and architecture, but no two studies investigated the same ECM feature. Of note, DiFrancesco *et al.* (1995) studied several different ECM features (calcification, collagen fibre organisation, elastic fibres, and GAG/proteoglycan content) in parallel[27], providing an overview of this tissue in OA.

#### 3.2.2 Fat pad in human OA

Two studies were identified for infrapatellar fat pad[30, 31]. Grevenstein *et al.* (2020) showed no change in COMP content between OA and control fat pads[30] and Belluzzi *et al.* (2020) showed that there was a decrease in collagen I and III expression in OA fat pads compared to controls [31].

#### 3.2.3 Intervertebral disc in human OA

One study was identified for intervertebral disc. Cheng *et al.* (1996) showed an increase in calcification with increasing OA grade in intervertebral discs[32].

#### 3.2.4 Ligaments in human OA

Out of the 14 studies on ligaments, 12 studies focussed on ACL and/or PCL of the knee[33–44], and two studies looked at ligaments in the thumb, one covering the palmar beak ligament and one covering volar anterior oblique (AOL) and dorsoradial (DRL) ligaments[45, 46]. Studies mostly focussed on collagen fibre organisation, which generally decreased in OA compared to control[38, 39, 41]. Studies without controls report disorganised and irregular collagen fibre organisation in OA ligaments. Other identified studies confirmed the presence of aggrecan, collagens I, II, and III, and decorin, suggest an increase in calcification and GAG/proteoglycan content, and no change in overall collagen content.

#### 3.2.5 Meniscus in human OA

Studies on human meniscus (n = 41) covered a wide range of ECM components, architectural changes, as well as measurements of viscoelastic properties[47–87]. Most studies concur on an increase in calcification and GAG/proteoglycan content, and consistently show a decrease in collagen fibre diameter and organisation. The presence or change in many other ECM components has been studied, including aggrecan, biglycan, cartilage intermediate layer protein, collagens and collagen crosslinks, COMP, decorin, fibromodulin, GAG components, hydroxyproline, keratocan, lubricin, and lumican. Most notable, all proteomics studies included in this systematic review evaluated human OA meniscus, identifying a range of ECM and ECM-associated proteins[52, 76, 79]. Two of these studies (Folkesson *et al.* (2020) and Roller *et al. (*2015)) also analysed control samples and found several proteins to be changed in OA compared to control tissue. For example, both studies report an increase in collagen type VI alpha 1 and collagen type VI alpha 2 in OA, and Folkesson *et al.* (2020) found a change in protein abundance in several small leucin-rich proteoglycans, such as an increase lumican and decrease in decorin, an increase in the proteoglycans aggrecan and versican, and a decrease in collagens type III and V [52, 79]. Finally, the results on viscoelastic properties are conflicting: whereas some studies show an increase in elastic modulus [66] and instantaneous modulus[77], another study showed a decrease in these moduli [47].

#### 3.2.6 Skeletal muscle in human OA

All four studies on human skeletal muscle studied the ECM components in the vastus medialis or vastus lateralis of the quadriceps muscle[88–91]. These studies demonstrated the presence [89] or increase [91] in collagens type I, III, and IV compared to control. In addition, these studies show the presence of calcification and laminin[88, 91], and an increase in collagen and GAG content[90].

#### 3.2.7 Synovium in human OA

Synovial tissue was studied in several joints, including 17 studies on the knee[68, 92–107], 5 studies on hip[108–112], 6 studies used a combination of knee and hip synovium [113–118], 2 studies on TMJ[119, 120], and 10 studies did not report the joint site[121–130]. The ECM components that were most often studied in human synovium were collagens, fibronectins, and laminins. Other ECM features covered by the included studies are calcification, collagen content, collagen fibre organisation, collagen cross-links, COMP, elastin, fibromodulin, GAG components, latent TGF- β - binding protein 1, lumican, reticulin, and vitronectin. While the presence and tissue distribution of these components has been clearly shown by several studies, the changes between OA and normal tissue remains unclear with most studies lacking healthy control groups; instead, OA is often the comparator group in studies investigating rheumatoid arthritis.

#### 3.2.8 Tendon in human OA

Human tendon studies covered a range of different tendons across the body, including Achilles, (long head of) biceps, subscapularis, gluteus medius, and internal obturator[131–135]. Discordant results between studies of anatomically distinct tendons are unsurprising, but disagreement was also seen for two studies on biceps tendon. For example, GAG/proteoglycan content was increased in the long head of biceps and internal obturator tendon[134, 135], unchanged in another study on biceps tendon and subscapularis tendon[132], and decreased in gluteus medius tendon in OA compared to control[133]. Similarly, increased calcification was seen in obturator tendon[135], while there was no difference in subscapularis, and a decrease in biceps tendon[132]. In terms of architecture, 3 out of 4 studies reporting on collagen fibre organisation report a decrease in organisation[132, 133, 135], while the last reported no difference compared to control[134]. An increase in collagen fibre diameter was found in internal obturator and biceps tendon[132, 135], while no difference was seen in subscapularis and gluteus medius tendons[132, 133]. Finally, no difference was found in the protein expression of collagen I and II and decorin[134].

### 3.3 Animal studies

Animal studies followed a similar pattern as human studies regarding the most studied tissues: synovium (n = 15), meniscus (n = 12), ligament (n = 6), skeletal muscle (n = 1), tendon (n = 1), and capsule (n = 1) (Supplementary Table 2). A broad range of species, strains, and models were used, all looking at the stifle joint of these animals. Overall, these studies generally found increases in ECM components such as collagen and disrupted ECM architecture, including a decrease in collagen fibre organisation in most tissues (Table 2). Viscoelastic properties were mainly studied in meniscus, where the elastic and instantaneous modulus tended to decrease.

**Table 2.**
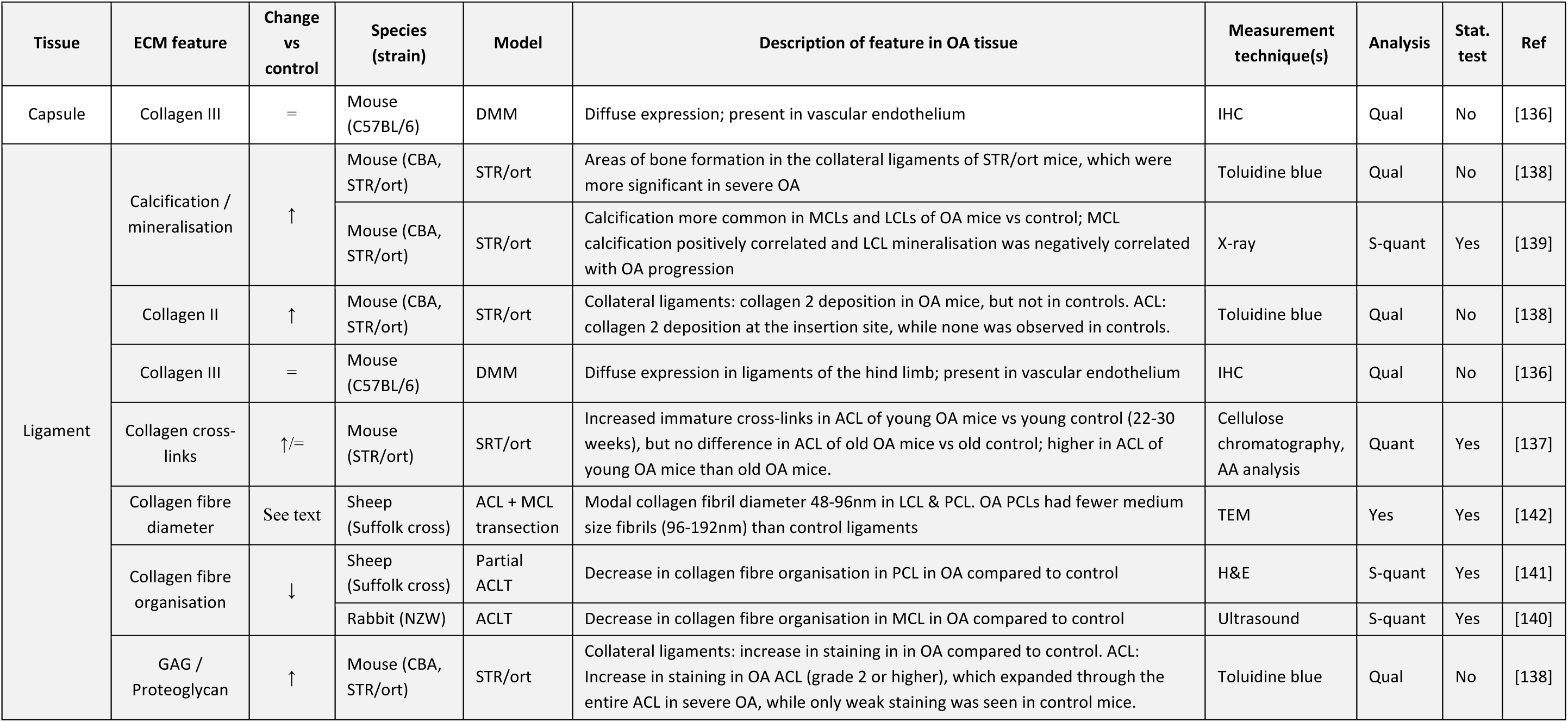
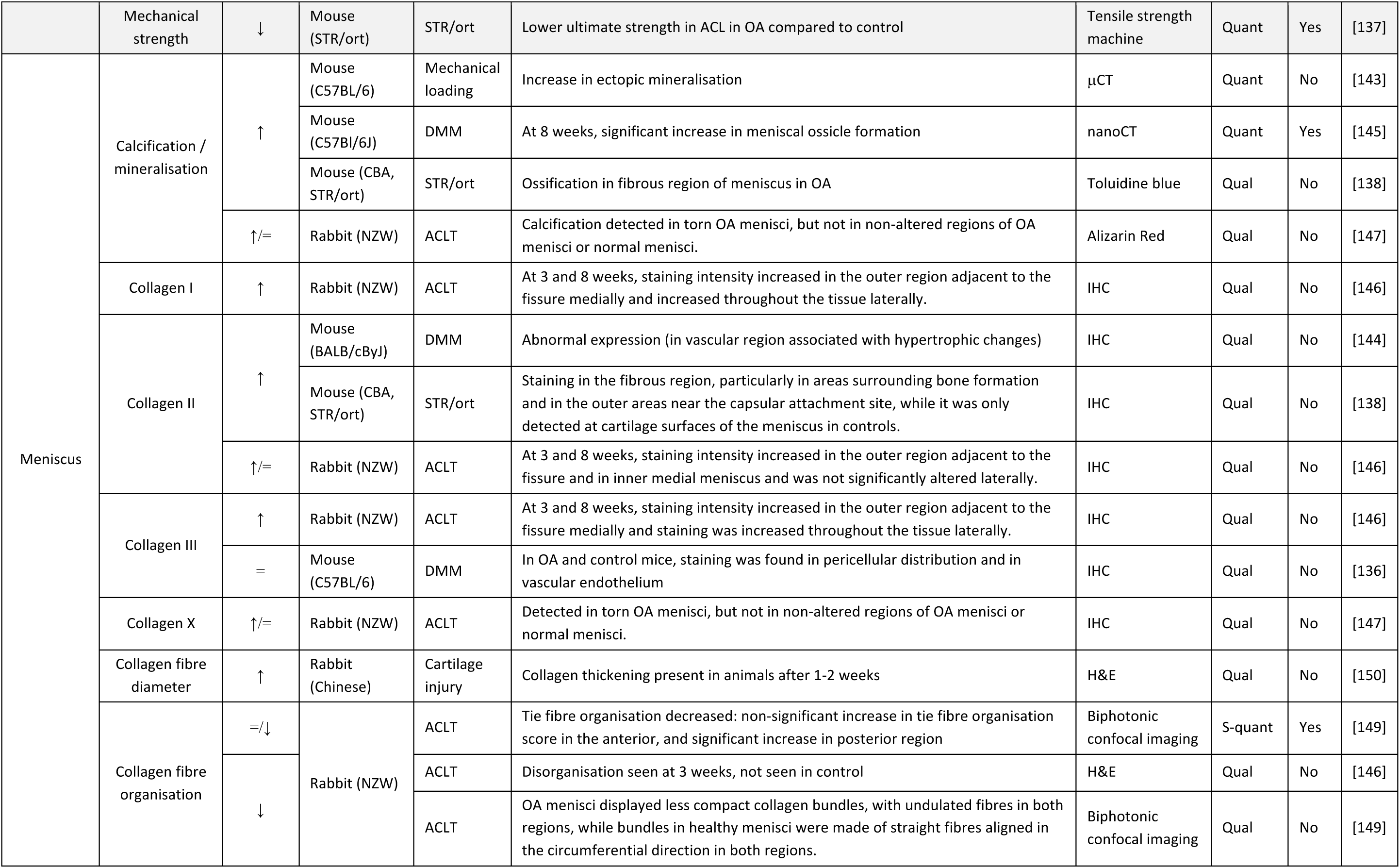
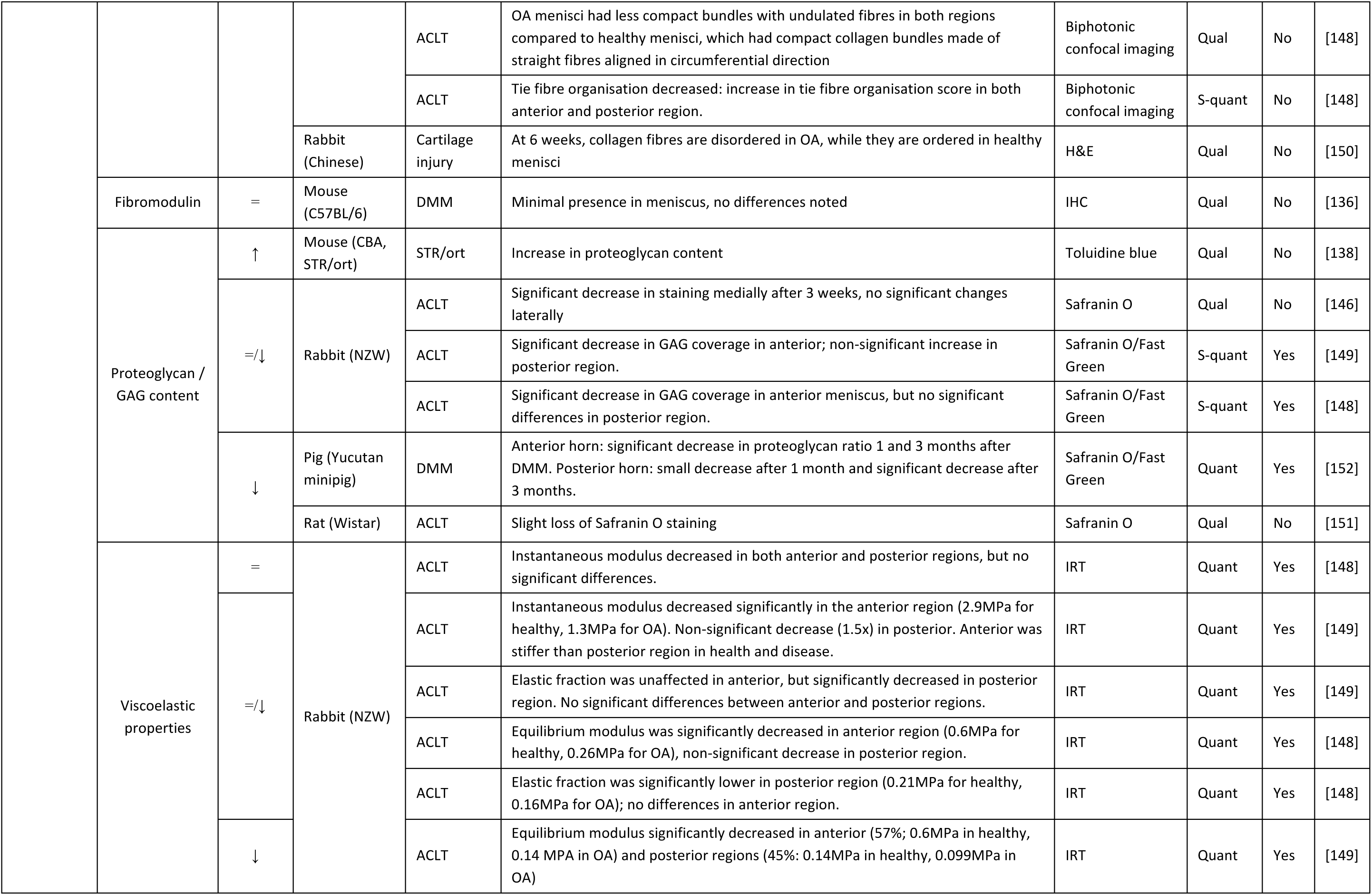
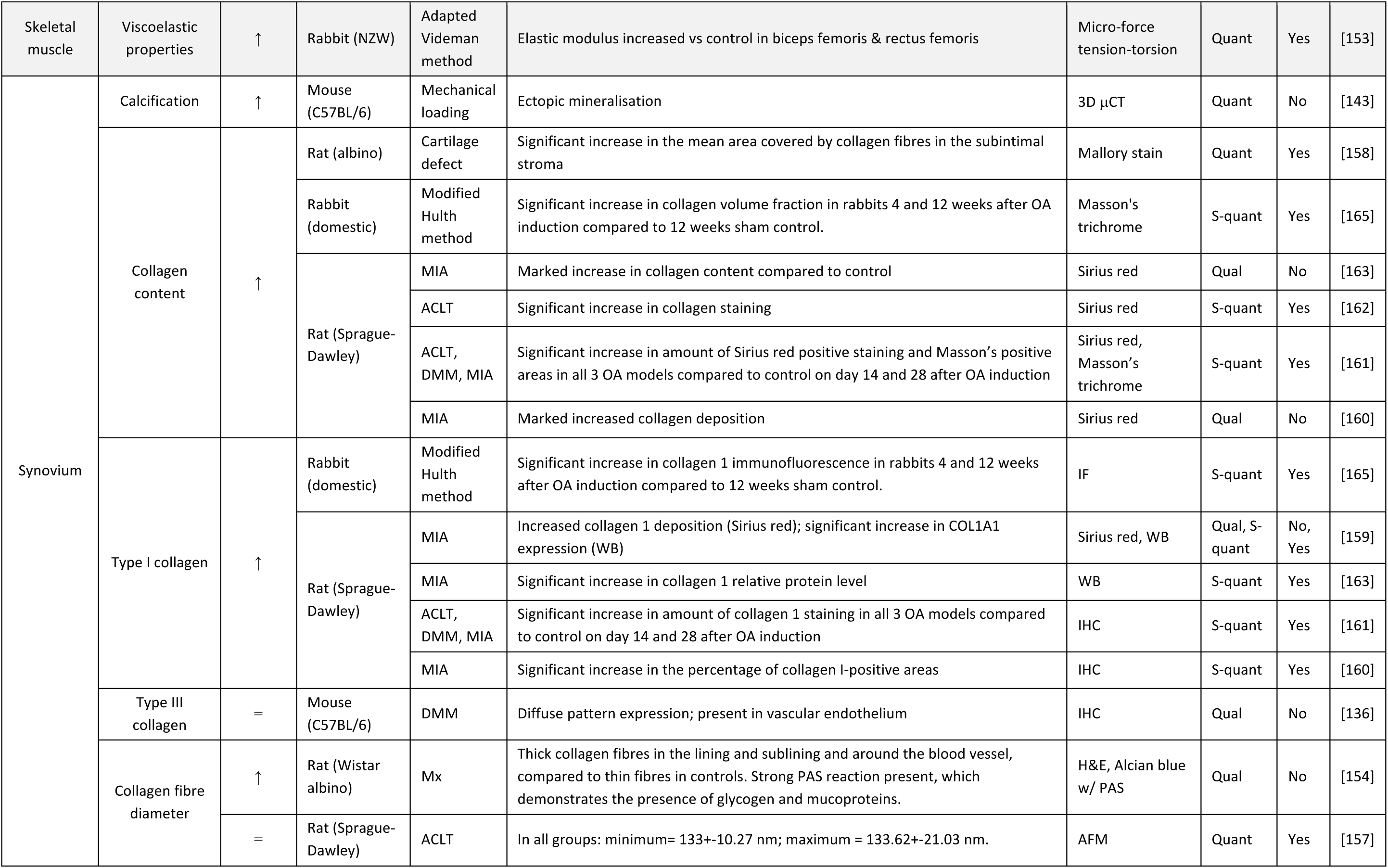
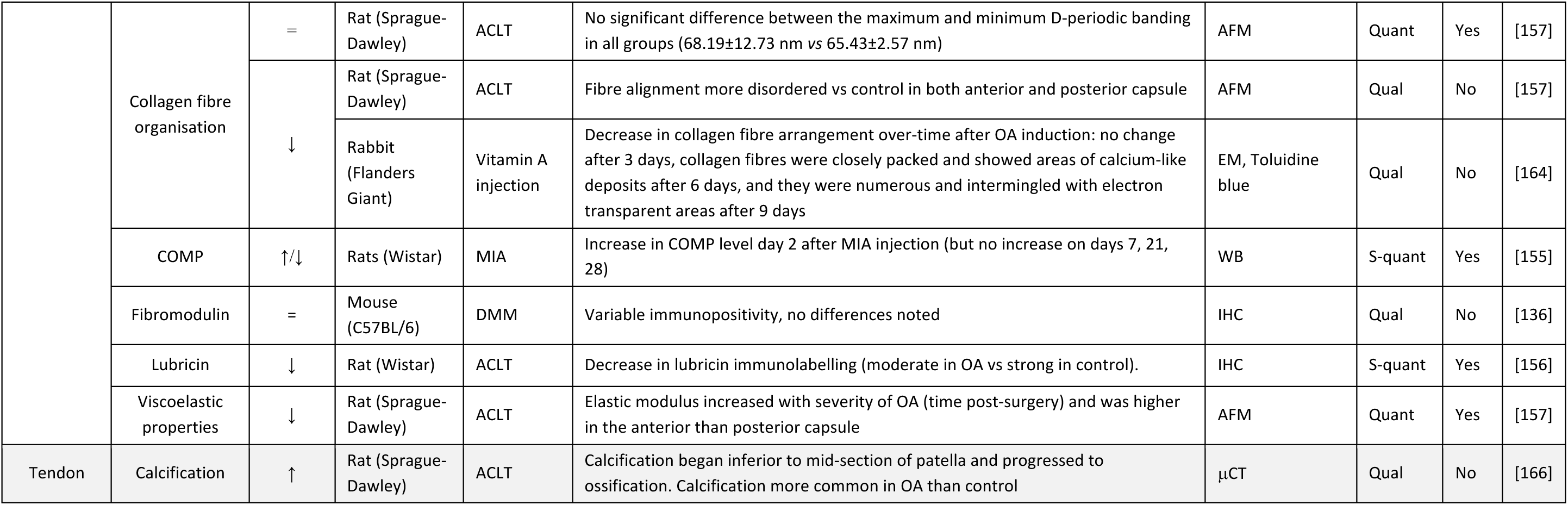
Structural ECM composition and architecture in animal models of OA. The studied joint in all studies is the stifle joint. Abbreviations: AA = amino acid, ACL = anterior cruciate ligament, ACLT = anterior cruciate ligament transection, AFM = atomic force microscopy, CT = computed tomography, DMM = destabilisation of the medial meniscus, ECM = extracellular matrix, EM = electron microscopy, GAG = glycosaminoglycan, H&E = haematoxylin and eosin, IHC = immunohistochemistry, IRT = indentation relaxation tests, LCL = lateral cruciate ligament, MCL = medial cruciate ligament, MIA = monoiodoacetate, Mx = meniscectomy, N/A = not applicable, NR = not reported, NZW = New Zealand white, OA = osteoarthritis, PAS = periodic acid Schiff, PCL = posterior cruciate ligament, qual = qualitative, quant = quantitative, s-quant = semi-quantitative, WB = western blot.

#### 3.3.1 Capsule in animal models of OA

Only one study was identified on capsule. Loeser *et al.* (2013) studied capsule in the DMM model in CD57BL/6 mice[136]. Collagen III was found to be diffusely expressed in OA capsule, and mostly present in vascular endothelium. Interestingly, this study also assessed the meniscus, ligament, and synovium, taking a whole-joint approach to OA; they report a diffuse distribution of collagen type III similar to capsule in ligaments and synovium, while there was a pericellular distribution in meniscus.

#### 3.3.2 Ligament in animal models of OA

Ligaments were studied in animal models of OA in 4 studies using mouse models[136–139], 1 study using a rabbit model[140], and 2 studies using sheep models[141, 142]. A decrease in collagen fibre organisation was reported by two studies[140, 141]. All other reported ECM features were only present in one study. These features include calcification, mineralisation, collagen II and III, collagen cross-links, collagen fibre diameter, GAG/proteoglycan content, and mechanical strength.

#### 3.3.3. Meniscus in animal models of OA

ECM changes in meniscus in animal models of OA were investigated by 5 studies using mouse models [136, 138, 143–145], 5 studies using rabbit models [146–150], 1 study using a rat model[151], and 1 study using a pig model[152]. Overall, these studies show an increase in calcification and collagen types I, II, III, and X, and a decrease in collagen fibre organisation. Most studies show a decrease in GAG/proteoglycan content and viscoelastic properties in at least parts of the meniscus. In addition, thickening of the collagen fibres and no change in fibromodulin were found.

#### 3.3.4 Skeletal muscle in animal models of OA

One study was identified that investigated skeletal muscle. Shi *et al.* (2020) studied the elastic modulus in biceps femoris and rectus femoris muscles in an adapted Videman method in rabbits; they report an increase in elastic modulus in OA compared to control[153].

#### 3.3.5 Synovium in animal models of OA

Synovium was investigated in 2 studies using mouse models[136, 143], 10 studies using rat models[154–163], and 2 studies using rabbit models[164, 165], and 1 study using a sheep model of OA[141]. All studies on calcification, collagen content, and collagen I showed an increase in OA compared to control. However, results on collagen fibre organisation and collagen fibre diameter were less clear, with some studies reporting no change, while others reported a decrease in collagen fibre organisation and increase in collagen fibre diameter. Other studied features included type III collagen, COMP, fibromodulin, lubricin, and viscoelastic properties (elastic modulus), which were all only reported on by a single study.

#### 3.3.6 Tendon in animal models of OA

Tendon was investigated in one study by McErlain *et al.* (2008) using an ACL transection model in rats. They found calcification of the patellar tendon to be more common in OA than control animals[166].

### 3.4 Bias analysis

The risk of bias varied between studies but was generally high (Supplementary Table 3). The potential for confounding bias was common, with many human studies failing to report on the age, sex and BMI of participants. Frequently, OA diagnoses were stated without reference to the diagnostic criteria used. Most studies failed to report on the blinding of assessors, even when qualitative histological observations were made. Purely qualitative observations were common, although semi-quantitative scoring systems were increasingly used in more recent studies. However, many quantitative and semi-quantitative differences between healthy and osteoarthritic tissues were not statistically analysed.

## 4 Discussion

Despite OA becoming more widely accepted as a whole joint disease, the role of and the changes to non-cartilage soft joint tissues remain underexplored. This study aimed to collate current knowledge on the structural ECM of these tissues to summarise and highlight gaps in existing knowledge. For instance, tissues such as the joint capsule and fat pad are very poorly defined, perhaps reflecting their perceived importance in OA. Overall, the studies included in this review show that the expression of many structural ECM components changes in disease, within an ECM that becomes less organised with increasing joint degeneration.

Human studies covered a range of tissues and ECM features, but focused mainly on calcification, the expression of proteoglycans, and the expression, fibre diameter, and fibre organisation of collagens. While recent studies begin to define the presence and distribution of many ECM components, a frequent lack of well-defined controls limit understanding of the changes in disease. Most ECM features are only described by one or a few studies, highlighting the broader studies on the subject. While studies that did look at the same ECM feature mostly agreed, this was not always the case. This included studies with control groups that investigated the collagen content in meniscus[78, 83], elastic modulus in meniscus[47, 66], chondroitin sulphate in synovium[102, 118], and calcification and GAG/proteoglycan content in tendon[132–135], which all contradict each other in terms of the direction of change. The summary and results tables highlight several potential factors for these differences already, including different analysis methods, differences in tissue joint origin, and differences between the microanatomical area of tissue studied. This emphasizes the importance of in-depth reporting of tissue metadata and methods.

Several recent human studies, mostly in ligaments, tendon, and meniscus, have begun to study both compositional and architectural ECM features within a single tissue. Importantly, such studies can begin to dissect the relationship, including causality, between changes in ECM composition, ECM architecture, and viscoelastic properties. For example, calcification of tendon has been shown to change its viscoelastic properties[167], while the mechanical properties of fibril-forming collagens are dependent on covalent cross-linking[168], and different matrix proteoglycans differ in their effects on cell-mediated collagen reorganisation[169].

Whole tissue proteomics, which can be used to study the ECM composition of a tissue holistically, was performed in three studies which all investigated OA meniscus[52, 76, 79]. While the study of ECM proteins using proteomic techniques are subject to methodological biases due to the fact that ECM proteins are large, heavily post-translationally modified, and highly insoluble[170], they are a powerful tool to better understand overall tissue composition and formulate new research questions. The application of this technique to other osteoarthritic tissues is likely to provide important insights.

In animal models, OA is induced in a range of species using varied surgical techniques and pharmacological interventions, with no animal model truly replicating human disease[171, 172]. Joint mechanics, inflammatory responses, and disease chronicity all vary between animal models [172, 173]. If ECM remodelling also differs between species and procedures, it can be assumed that not all animal models are equally suited to the study of changes in osteoarthritic ECM. Certain models may be generally more representative of changes seen in human OA, or better suited to the study of particular joint tissues or ECM features. This review covers a range of ECM changes in several different musculoskeletal soft tissues across different species and models. Although limited animal studies were eligible for inclusion in this review, some changes in ECM features could be compared between human OA and animal models. Generally similar trends could be seen as in humans, including a decrease in collagen fibre organisation and an increase in calcification across ligaments, meniscus, and synovium. However, other observations seem to contradict those in humans; for example, the decreased expression of collagens in human osteoarthritic menisci [54, 83, 86] is not reflected in data from any animal models in this review [138, 144, 146]. Therefore, the models used by these studies, namely the mouse STR/ort, rabbit ACLT, and mouse DMM models, respectively, might not be suitable to infer OA-related changes in human menisci. These results emphasize that more studies on ECM changes in non-cartilage soft-joint tissues in human OA and animal models must be compared before the validity of the latter can be accurately defined.

The strength of any systematic review is partly contingent on the quality of included studies. As discussed in section 3.4, the methodology of many studies conferred a high risk of bias, resulting in a low confidence in the evidence provided. In basic science studies utilising human samples, the baseline characteristics and clinical characterisation of OA patients are often missing, or lack necessary detail. Clinical background is a particularly important consideration in the context of soft tissue calcification, given that crystal depositional diseases, such as pseudogout, can drive OA pathology[174]. Patients’ clinical background is poorly reported throughout the literature, as is disease severity, despite ECM and other tissue components differing more from the physiological state with OA progression[38]. Although the search strategy covered many non-cartilage soft joint tissues, some tissues, such as the temporomandibular joint disc and acetabular labrum, were not included. In addition, the focus of this review was on structural components of the ECM, which are the elements that are studied most extensively and make up the majority of tissue ECM. However, this does mean that this work does not provide a complete account of all OA ECM, as non-structural matrix elements such as matricellular proteins or neoepitopes have not been reported on. Finally, a limitation of the review process is the data extraction, which was not done by two independent reviewers, but rather extracted by one reviewer and verified by the other reviewer. However, the effect of this is likely limited as a previous study has reported that while extraction by two independent reviewers is preferable, extraction by one reviewer with verification by a second reviewer has limited influence on the conclusions of a systematic review, especially considering a meta-analysis was not performed in the current work[175].

In the process of consolidating the current literature on this topic, this work highlights several practical and methodological challenges that have limited advances in knowledge of the understanding of structural ECM components, architectural features, and viscoelastic properties in non-cartilage soft tissues in OA. One of these problems, is the cross-sectional nature of studies. Cross-sectional studies are common across the OA field as tissues are only accessible at the time of joint replacement. Since OA can take decades to progress, the study of end-stage or advanced OA might not be very informative of the processes that are driving these changes. In addition, the lack of a healthy, or non-OA, comparator group in combination with the fact that many studies only report qualitative results, vastly reduces the depth of knowledge that can be gained from these studies. Finally, while many screened human and animal studies investigated both cartilage and other soft joint tissues, ECM is often studied exclusively in cartilage, with other features, such as cellularity and inflammatory markers being the focus in other tissues. This shows that while there is access to both the tissues and the methods to study ECM changes in non-cartilage soft tissues, their analysis is not seen as a priority. However, due to the limited definition of ECM in these tissues and their unknown contribution to disease development and progression, it is also possible that it remains unclear which ECM features should be focused on. Structural ECM encompasses a wide range of features that can be investigated with a plethora of different methods. To evaluate the most critical ECM features and applicable methods, studies investigating multiple ECM features in non-cartilage soft tissues across different stages of disease are required.

Recent studies have started to highlight the importance of ECM as a determinant of tissue architecture and cell behaviour in disease. For example, a recent review has highlighted the changes in microenvironment in rheumatoid arthritis synovium that occur in early in the development of the disease, which form important extracellular cues that shape the pathogenic cell behaviour during the onset and progression of disease[176]. Therefore, they argue that understanding the ECM changes across different tissues in a particular disease might not only be able to help with disease classification and patient stratification but could also hold promise for the development of treatments that target ECM[176]. These treatments might not only be able to modify pathogenic cell behaviour that could be driving the disease, but also impact on joint stiffness, which is one of the most common symptoms of OA[177]. All in all, more research is needed to unravel the presence and distribution of different ECM components and architectural features in joint tissues in health and in (different stages of) OA and interplay with tissue-resident and tissue-infiltrating cells. Future research will also help to differentiate between the remodelling process in different joint tissues, which contain unique cell populations and are exposed to different mechanical and inflammatory stimuli in OA. ECM remodelling may also differ between synovial joints, given their varied anatomical locations, mechanical functions, and the presence of joint-specific tissues such as menisci. Potential variation in pathophysiology between osteoarthritic joints has received little attention, with the predominance of studies on knee OA likely due to high disease prevalence in this joint and tissue being relatively accessible during commonly performed knee replacements. Therefore, the future of this field is both dependent on the thorough investigation of ECM features in soft joint tissues across multiple OA joints and varied stages of disease progression, as well as the rigorous reporting of patient characteristics of all tissue donors.

In conclusion, this systematic review gives an overview of the current knowledge of the presence and distribution of structural ECM components, as well as the changes in ECM components and architecture that occur throughout the osteoarthritic joint. Overall, the studies included in this review show that the expression of many structural ECM components changes in disease and that the ECM architecture becomes more disorganised with increasing joint degeneration. Furthermore, this review highlights practical and methodological challenges that have limited progress in the understanding of changes in ECM composition and architecture in non-cartilage soft tissues during OA development and progression, including the fact that historically studies have mainly focussed on other aspects of the disease in these tissues, such as inflammation. Given the role of ECM in influencing cell behaviour, further research to better understand the broad context within which cartilage is damaged in OA may enable a better understanding of the disease as well as potential treatments.

## Acknowledgements

We would like to thank Oxford University medical librarian Eli Harriss for her support in generating and executing the search strategy. We would like to thank Dr Mathew Baldwin for his helpful feedback on the design of this study.

## Author contributions

Conceptualization, J.Y.M. and S.J.B.S.; Formal analysis, J.Y.M. and I.G.A.R.; Investigation, J.Y.M. and I.G.A.R.; Data curation, J.Y.M. and I.G.A.R.; Writing - original draft, J.Y.M. and I.G.A.R.; Writing - review & editing, S.J.B.S; Project Administration, J.Y.M. and S.J.B.S.

## Data availability statement

The authors confirm that the data supporting the findings of this study are available within the article and its supplementary materials. In addition, the raw data from the data extraction process, which was used to populate Tables 1 and 2 and Supplementary Tables 1 and 2, is available upon reasonable request from the corresponding author (JYM).

