## Supplementary for "Defining the extracellular matrix in non-cartilage soft tissues in osteoarthritis – a systematic review"

**Supplementary Table 1.** Study characteristics of included human studies.

**Abbreviations**: ACL = anterior cruciate ligament, IQR = interquartile range, KCS = Knee Society Clinical Grading System, KL-grade = Kellgren-Lawrence grade, Med = median, NR = not reported, OA = osteoarthritis, OARSI = Osteoarthritis Research Society International, OCS = Outerbridge Classification System, PCL = posterior cruciate ligament, TKR = total knee replacement.

* = note on the control group of Herbert (1973): although 15 participants were in the control group in total, results in this review are based on the included adults only (11 participants).

### = note on the control group of Karube (1981): although non-adult participants were included in the study, the results in this review are based exclusively on the age groups with adults only.

| First author (year) | Country | Joint | Tissue (details) | Stage of disease / OA severity | Control population | No. of participants | Female (%) | Age (mean±SD (range)) | BMI (mean) | Ref |
|---|---|---|---|---|---|---|---|---|---|---|
| Abdul Sahib (2017) | Iraq | Knee | Ligament (ACL & PCL) | NR | No | OA: 50 | 70 | NR | NR | [33] |
| Abraham (2014) | USA | Knee | Meniscus (enthesis) | Advanced | Healthy tissue donors | OA: 7<br>Control: 8 | NR<br>NR | NR<br>55 (41-61) | NR<br>NR | [47] |
| Akisue (2002) | Japan | Knee | Ligament (PCL) | Ahlbäck grades 2-5 | Cadavers w/o knee OA | OA: 24<br>Control: 4 | 38<br>50 | 64.2 (40-84)<br>87 (78-98) | NR<br>NR | [34] |
| Allain (2001) | France | Knee | Ligament (ACL & PCL) | TKR | No | OA: 45 | 67 | 76 (68-84) | NR | [35] |
| Atik (2016) | Turkey | Knee | Meniscus (medial) | Medial arthroplasty | No | OA: 12 | 100 | 64 (59-71) | NR | [48] |
| Battistelli (2019) | Italy | Knee | Meniscus | KL grade 3-4 | Multi-organ donors w/o history of joint disorders | OA: 3<br>Controls: 3 | 33<br>33 | Med 72 (IQR 72-73.5)<br>Med 66 (IQR 63-69) | NR<br>NR | [49] |
| Belluzzi (2020) | Italy | Knee | Fat pad (infrapatellar) | End-stage | ACL reconstruction, med 8 months post-injury | OA: 25<br>Control: 28 | 72<br>25 | Med 68 (IQR 62-75)<br>Med 31 (22-42) | Med: 29.5<br>Med: 23.0 | [31] |
| Cameron (1973) | Canada | Hip | Capsule | End-stage | Not reported | OA: 25<br>Control: 15 | NR<br>NR | NR<br>NR | NR<br>NR | [25] |
| Campbell (2015) | Canada | Knee | Capsule | Mean KL grade >3 | No | OA: 21 | 57 | Mean >65 | NR | [26] |

| | | | | | | | | | | |
|---|---|---|---|---|---|---|---|---|---|---|
| Chang (2005) | Japan | NR | Synovium | Arthroplasty | No | OA: 5 | NR | NR | NR | [121] |
| Cheng (1996) | Belgium | Spine | Intervertebral disc | KL grade 1-4 | Cadavers w/o spinal OA | OA: 51<br>Control: 9 | 50 | 67.3 (23-87) | NR | [32] |
| Christensen (2019) | Denmark | Hip | Synovium | Advanced | No | OA: 6 | 3 | Med 66.5 (IQR 62–69) | Med 30.8 | [108] |
| Cillero-Pastor (2015) | Netherlands | Knee | Synovium | TKR | Adult tissue donors | OA: 3<br>Control: 3 | NR<br>NR | 69-82<br>60-78 | NR<br>NR | [92] |
| Cutolo (1992) | Italy | Knee | Synovium | End-stage | Knee trauma | OA: 4<br>Control: 4 | NR<br>NR | NR<br>NR | NR<br>NR | [93] |
| Dessombz (2013) | France | Knee | Meniscus (medial) | KL mean grade 4 | No | OA: 6 | 83 | 74 (SD 9) | 28 | [50] |
| DiCesare (1999) | USA | Knee | Synovium | Knee replacement | No | OA: 3 | NR | NR | NR | [94] |
| DiFrancesco (1995) | USA | Hip | Capsule | Advanced | No | OA: 109 | 57 | 67.1 (39-92) | NR | [27] |
| Dijkgraaf (1997) | Netherlands | TMJ | Synovium | Early to advanced | No | OA: 31 | 81 | 30.1 | NR | [119] |
| Doerschuk (1999) | USA | Thumb | Ligament (palmar beak) | Mankin grades 3-14 | No | OA: 18 | 72 | NR | NR | [45] |
| Ea (2013) | France | Knee | Synovium | KL grades 0-3 | No | Total: 31 | NR | NR | NR | [95] |
| Ene (2015) | Romania | Knee | Synovium | Early and late stage | No | OA: 43 | 77 | 49-76 | NR | [96] |
| Exposito Molinero (2016) | Spain | NR | Tendon (Achilles) | NR | No | OA: 24 | 46 | 50.7 | NR | [131] |
| Fan (2012) | China | Knee | Synovium | NR | No | OA: 6 | 50 | Med 60 (range 48-77) | NR | [97] |
| Fink (2007) | Germany | Knee | Skeletal muscle (vastus medialis) | KL grade 4 | No | OA: 78 | 79 | 68.5 (51-83) | <20: 3<br>20-24: 12<br>25-29: 31<br>>29: 32 | [88] |
| Fischenich (2015) | USA | Knee | Meniscus (medial & lateral) | TKR | No | OA: 24 | 54 | 57.8 ± 4.6 | 31.9 | [51] |
| Folkesson (2020) | Sweden | Knee | Meniscus (posterior | OCS 4 medially; | Cadavers (w/o joint disease) | OA: 9<br>Control: 10 | 56<br>50 | 62.0 (50-75)<br>51.4 (18-77) | 29.0<br>28.0 | [52] |

| | | | | | | | | | | |
|---|---|---|---|---|---|---|---|---|---|---|
| | | | horns of medial) | OCS 0-1 laterally | | | | | | |
| Fuhrmann (2015) | Germany | Knee | Meniscus | TKR | No | OA: 9 | 67 | 69.6 (56-80) | NR | [53] |
| Ghosh (1975) | Australia | Knee | Meniscus | NR | No | OA: 5 | NR | NR | NR | [54] |
| Grevenstein (2020) | Germany | Knee | Fat pad (infrapatellar) | TKR | Trauma (ACL rupture) | OA: 14<br>Control: 11 | 50<br>27 | 63.8 ± 17.6<br>33.7 ± 14.8 | NR | [30] |
| Haut Donahue (2021) | USA | Knee | Meniscus (medial & lateral enthesis) | TKR | No | OA: 7 | NR | 40-75 | NR | [55] |
| Heinegård (1968) | Sweden | Knee | Capsule | NR | No | OA: 8 | 88 | 60-80 | NR | [28] |
| Herbert (1973) | UK | Hip | Capsule | NR | Cadavers w/o joint disease | OA: 12<br>Control: 11* | NR<br>NR | 30-70<br>NR | NR<br>NR | [29] |
| Hino (1995) | Japan | NR | Synovium | NR | No | OA: 4 | NR | NR | NR | [122] |
| Hino (2020) | Japan | Knee | Meniscus (posterior root) | NR | No | OA: 7 | 57 | 75 (67-86) | NR | [56] |
| Ibrahim (2019) | Sweden | Shoulder | Tendon (biceps & subscapularis | Total shoulder arthroplasty | Proximal humeral fracture | OA: 13<br>Control: 13 | NR<br>NR | Med 67<br>Med 70 | NR<br>NR | [132] |
| Ibrahim (2021) | Sweden | Hip | Tendon (Gluteus Medius) | Total hip arthroplasty | Femoral head fracture | OA: 29<br>Control: 25 | 66<br>76 | 70<br>73 | NR<br>NR | [133] |
| Ishizuka (2016) | Japan | Knee | Meniscus | TKR | No | OA: 30 | 57 | 71 | NR | [57] |
| Itokazu (1998) | Japan | NR | Synovium | NR | Traumatic injury | OA: 22<br>Control: 16 | 41<br>25 | 59<br>21 | NR<br>NR | [123] |
| Jacquet (2018) | France | Knee | Meniscus | TKR | No | OA: 5 | 40 | 64 | 23.2 | [58] |
| Jacquet (2019) | France | Knee | Meniscus (lateral) | Advanced medially, KL grade <2 laterally | No | OA: 24 | 50 | 64 | NR | [59] |

| | | | | | | | | | | |
|---|---|---|---|---|---|---|---|---|---|---|
| Johnson (2001) | USA | Knee | Meniscus (medial) | Advanced (joint replacement) | Cadavers | OA: 5<br>Control: 5 | NR<br>NR | NR<br>NR | NR<br>NR | [60] |
| Karjalainen (2021) | Finland | Knee | Meniscus (medial & lateral) | Outerbridge IV medially; 0/I medially | Cadavers | OA: 10<br>Control: 10 | 50<br>50 | 63 ± 7<br>51 ± 17 | NR | [61] |
| Karube (1981) | Japan | Knee | Meniscus | NR | NR | OA: 3<br>Control: 17[#] | NR | 48-62<br>13-62[#] | NR<br>NR | [62] |
| Katsuragawa (2010) | Japan | Knee | Meniscus (ant. horn and body of medial and lateral) | End-stage | Cadavers w/o joint disease | OA: 40<br>Control: 22 | NR<br>NR | NR<br>84 (73-92) | NR<br>NR | [63] |
| Kaufmann (2003) | Germany | Knee | Synovium | Knee replacement | No | OA: 21 | 76 | 71 (49-82) | NR | [98] |
| Kiraly (2017) | USA | Knee | Meniscus (posterior medial) | TKR | No | OA: 10 | NR | NR | NR | [64] |
| Klareskog (1986) | Sweden | NR | Synovium | Arthroplasty | No | OA: 15 | NR | NR | NR | [124] |
| Kodama (2018) | Japan | Knee | Meniscus (ant. & post. sections) | KL grade 4 (TKR) | No | OA: 26 menisci | 84 | 76 ± 6.7 (68-84) | NR | [65] |
| Komro (2020) | USA | Knee | Ligament (ACL) | OCS mean 3.6 (range 3.2-4.0) | Cadaver (normal macroscopic ACL) | OA: 6<br>Control: 16 | 100<br>50 | 66 ± 9.6<br>75 ± 12.3 | 37.6<br>23.0 | [36] |
| Konttinen (1999) | Finland | Hip | Synovium | NR | ACL tear (2), meniscal injury (3) | OA: 10<br>Control: 5 | 70<br>60 | 71 (41-84)<br>41 (22-53) | NR<br>NR | [109] |
| Konttinen (2001) | Finland | Hip | Synovium | THA | No | OA: 10 | 60 | 70 (40-83) | NR | [110] |
| Kragstrup (2019) | USA | Knee | Synovium | KL grade 2-4 | No | OA: 12 | NR | NR | NR | [99] |
| Kriegsman (2004) | Germany | NR | Synovium | NR | No | OA: 3 | NR | NR | NR | [125] |
| Kumagai (2012) | USA | Knee | Ligament (ACL & PCL) | Koshimoto's grade 3-5 | No | OA: 28 | 82 | 77 (67-84) | NR | [37] |

| | | | | | | | | | | |
|---|---|---|---|---|---|---|---|---|---|---|
| Kwok (2014) | USA | Knee | Meniscus | OARSI score 4 | Healthy donors (OARSI 0-1) | OA: 3<br>Control: 3 | NR<br>NR | 75 (61-90)<br>31 (20-41) | NR<br>NR | [66] |
| Levy (2013) | USA | Knee | Ligament (ACL & PCL) | OCS score 0-4 | Cadavers w/o joint degeneration | 65 (111 OA & 9 control knees) | 54 | 67 ± 19 (23-92) | 24.8 | [38] |
| Li (2000) | Finland | Hip | Synovium | Hip replacement | No | OA: 10 | NR | NR | NR | [111] |
| Lopez-Franco (2016) | Spain | Knee | Meniscus (medial) | Knee replacement | No | OA: 31 | 74 | 72 ± 6.7 (60-84) | NR | [67] |
| Mapp (1985) | UK | Knee & hip | Synovium | NR | No | OA: 4 | 100 | 60.8 (46-69) | NR | [113] |
| Marczak (2017) | Poland | Knee | Ligament (PCL) | Ahlbäck grade 3-5 | Cadavers w/o joint damage | OA: 50<br>Control: 10 | 80<br>70 | 71 (53-84)<br>72 (67-78) | NR<br>NR | [39] |
| Martins (2018) | Brazil | Knee | Ligament (PCL) | Ahlbäck grade 1-5 | No | OA: 85 | 81 | 70 (53-87) | NR | [40] |
| Masuda (1991) | Japan | Knee | Synovium, meniscus | NR | No | OA: 10 | NR | 76 | NR | [68] |
| Mattiello-Sverzut (2013) | Brazil | Knee | Skeletal muscle (vastus lateralis) | KL grade 2.3 ± 1.2 | No | OA: 6 | 33 | 62 ± 5 | 28.1 | [89] |
| Mazzocca (2013) | USA | Shoulder | Tendon (long head of biceps) | NR | Cadavers w/o joint damage | OA: 9<br>Control: 9 | 22<br>56 | 59 ± 11.6<br>58 ± 14.7 | NR<br>NR | [134] |
| McDaniel (2017) | USA | Knee | Meniscus (medial & lateral) | Knee replacement | Cadavers w/o joint damage | OA: 14<br>Control: 14 | 50<br>64 | 62 ± 7.7<br>67 ± 10.6 | 34.6<br>NR | [69] |
| Meknas (2012) | Norway | Hip Tendon | Tendon (internal obturator) | NR | Femoral fracture | OA: 10<br>Control: 10 | 60<br>80 | Med 60 (48-75)<br>Med 83 (60-90) | NR<br>NR | [135] |
| Melrose (2008) | Australia | Knee | Meniscus (medial & lateral) | Joint replacement | Cadavers w/o cartilage damage | OA: 17<br>Control: 6 | 11<br>NR | 77.8 ± 5.4 (70-88)<br>(60-75) | NR<br>NR | [70] |
| Mine (2013) | Japan | Knee | Meniscus | TKR | No | OA: 12 | NR | NR | NR | [71] |

| | | | | | | | | | | |
|---|---|---|---|---|---|---|---|---|---|---|
| Mobargha (2014) | Sweden | Thumb | Ligament (volar anterior oblique & dorsoradial) | Eaton stage 2-3 | No | OA: 11 | 91 | 67 (51-83) | NR | [46] |
| Monibi (2021) | USA | Knee | Meniscus (posterior medial) | Knee replacement | No | OA: 8 | 63 | 65 ± 5.5 | NR | [72] |
| Musumeci (2014) | Italy | Knee | Meniscus (medial & lateral) | KL grade 2-3 | ACL reconstruction | OA: 40<br>Control: 9 | 38<br>44 | Med 68 (51-76)<br>Med 60 (45-66) | NR<br>NR | [73] |
| Nagata (2000) | Japan | Knee | Meniscus (medial) | KCS score: mean 52.6 | No | OA: 11 | 73 | 79 (73-86) | NR | [74] |
| Nakahara (2013) | USA | Knee | Ligament (ACL) | KCS score 2-4 | Normal cadaver, aged controls (cartilage score 1) | OA: 16<br>Cadaver: 13<br>Aged: 8 | 50<br>Unclear<br>63 | 80 ± 11.4<br>37 ± 11.0<br>77 ± 12.9 | NR<br>NR<br>NR | [41] |
| Nakashima (1998) | Japan | Knee | Synovium | KCS score (2-4) | No | OA: 9 | 67 | 57 ± 8 (47-70) | NR | [100] |
| Nelissen (2001) | Netherlands | Knee | Ligament (PCL) | Ahlbäck grade 3-5 | No | OA: 11 | 82 | 75 (60-85) | NR | [42] |
| Nikkari (1995) | Finland | Knee | Synovium | Knee replacement | No | OA: 6 | NR | 63 | NR | [101] |
| Nishida (1995) | Japan | Knee | Synovium | Knee replacement | No | OA: 18 | 83 | 72 (62-86) | NR | [102] |
| Noehren (2018) | USA | Knee | Skeletal muscle (vastus lateralis) | KL grade 2-3 | KL grade 0-1 | OA: 24<br>Control: 15 | 42<br>67 | 60 ± 5.5 (52-73)<br>64 ± 6.9 (54-74) | 28.4<br>26.9 | [90] |
| Okamoto (2015) | Japan | TMJ | Synovium | NR | Cancer patient cadavers | OA: 5<br>Control: 3 | 60<br>67 | 60 (20-72)<br>64 (61-70) | NR<br>NR | [120] |
| Park (2015) | South Korea | Knee | Meniscus (medial posterior root) | KL grade 3-4 | Normal joint cadavers | OA: 99<br>Control: 3 | NR<br>33 | (50-85)<br>54 (47-61) | NR<br>NR | [75] |

| | | | | | | | | | | |
|---|---|---|---|---|---|---|---|---|---|---|
| Park (2021) | South Korea | Knee | Meniscus (medial & lateral) | Knee replacement | No | OA: 12 | 100 | 73 (65-82) | NR | [76] |
| Poduval (2010) | Finland | NR | Synovium | NR | Knee trauma | OA: 5<br>Control: 5 | NR<br>NR | NR<br>NR | NR<br>NR | [126] |
| Pollock (1990) | UK | Knee & hip | Synovium | NR | Trauma | OA: 7<br>Control: 4 | 71<br>50 | 69 (58-83)<br>38 (27-60) | NR<br>NR | [114] |
| Pordzik (2020) | Germany | Knee | Meniscus (medial & lateral) | KL grade 4 | NR | OA: 26<br>Control: 14 | 73<br>NR | Overall: 72 ± 6.7 | NR | [77] |
| Rafael (2014) | Portugal | Knee | Synovium | Knee replacement | Normal joint cadaver | OA: NR<br>Control: NR | NR<br>NR | NR<br>NR | NR<br>NR | [103] |
| Rajgopal (2014) | India | Knee | Ligament (PCL) | KCS mean 42 | No | OA: 62 | 74 | 67 ± 11 (49-91) | NR | [43] |
| Richardot (2009) | France | Knee | Synovium | Ahlbäck grade 3-5 | No | OA: 4 | 100 | (54-73) | NR | [104] |
| Rinaldi (1998) | Germany | Knee & hip | Synovium | NR | No | OA: 18 | NR | NR | NR | [115] |
| Roller (2015) | USA | Knee | Meniscus (medial & lateral) | KL grade mean 3.94 | Cadaver or amputation (KL 0) | OA: 23<br>Control: 5 | NR<br>NR | 60 (36-81)<br>77 (64-87) | NR<br>NR | [78] |
| Roller (2015) | USA | Knee | Meniscus | Mild to severe OA | Normal joint cadaver | OA: 6<br>Control: 3 | 50<br>NR | (44-69)<br>(64-78) | NR<br>NR | [79] |
| Saito (2002) | Japan | Knee | Synovium (medial & lateral) | Advanced disease | No | OA: 21 | NR | 64 (47-78) | NR | [105] |
| Santiago (2006) | Spain & France | NR | Synovium | NR | No | OA: 8 | NR | NR | NR | [127] |
| Schneider (1994) | Germany | NR | Synovium | NR | Meniscal lesions | OA: 8<br>Control: 31 | NR<br>NR | NR<br>NR | NR<br>NR | [128] |
| Scott (1984) | UK | NR | Synovium | NR | Meniscal lesions | OA: 10 | 60 | (37-72) | NR | [129] |
| Scott (1981) | UK | Hip & knee | Synovium | NR | No | OA: 8 | 63 | 61 (37-72) | NR | [116] |
| Serrao (2014) | Brazil | Knee | Skeletal muscle | KL grade 1-2 | Healthy controls | OA: 18<br>Control: 17 | NR<br>NR | 51 ± 6.3<br>52 ± 8.1 | 29.5<br>27.3 | [91] |

| | | | | | | | | | | |
|---|---|---|---|---|---|---|---|---|---|---|
| | | | (vastus lateralis) | | | | | | | |
| Sladojevic (2016) | Bosnia & Herzegovina | Knee | Meniscus (medial & lateral) | Advanced disease | No | OA: 35 | 86 | 69.3 (56-78) | 30.8 | [80] |
| Son (2013) | USA | Knee | Meniscus (medial & lateral) | TKR | No | OA: 14 | 57 | 65 | NR | [81] |
| Sun (2010) | USA | Knee | Meniscus (posterior medial) | End-stage disease | No | OA: 8 | 6 | 57 (42-70) | NR | [82] |
| Sun (2012) | USA | Knee | Meniscus (medial) | End-stage disease | No | OA: 8 | 6 | 57 (42-70) | NR | [83] |
| Takahashi (1996) | Japan | Knee | Synovium | TKR | No | OA: 10 | NR | 58-79 | NR | [106] |
| Takahashi (1998) | Japan | Knee | Meniscus (central & peripheral) | TKR | No | OA: 21 | NR | 69 (40-84) | NR | [84] |
| Turdean (2017) | Romania | Hip | Synovium | KL grade 3-4 | No | OA: 57 | 44 | 64 (24-83) | NR | [112] |
| Van Linthoudt (1997) | USA | Hip & knee | Synovium | KL grade 2-4, mean 3.25 | No | OA: 16 | 0 | 68 (54-76) | NR | [117] |
| Wang (2020) | UK | Knee | Meniscus (medial) | TKR | No | OA: 10 | 40 | 66 (46-87) | NR | [85] |
| Wang (2018) | China | Knee | Synovium | TKR | Lower limb amputation | OA: 31<br>Control: 5 | Overall: 20 | Overall: (48-69) | NR | [107] |
| Warnecke (2020) | Germany | Knee | Meniscus (lateral) | TKR | No | OA: 24 | 67 | 67 ± 9.0 | 29 | [86] |
| Worrall (1991) | UK | NR | Synovium | TKR | Malignancy | OA: 8<br>Control: 5 | 63<br>NR | 62<br>NR | NR<br>NR | [130] |
| Worrall (1994) | UK | Knee & hip | Synovium | Replacement arthroplasty | (ankle, knee, elbow) | OA: 6<br>Control: 6 | NR<br>NR | NR<br>NR | NR<br>NR | [118] |
| Zhang (2012) | USA | Knee | Meniscus | NR | No | OA: 18 | NR | (50-81) | NR | [87] |
| Zhu (2017) | China | Knee | Ligament (ACL) | TKR | No | OA: 20 | NR | 65 ± 5.6 | NR | [44] |

**Supplementary Table 2.** Study characteristics of animal studies. Joint of interest is the knee joint, unless otherwise specified. Age/time is reported in days (d), weeks (w), months (m), or years (y).

**Abbreviations**: ACL = anterior cruciate ligament, ACLT = anterior cruciate ligament transection, DJD = degenerative joint disease,DMM = destabilisation of the medial meniscus, LCL = lateral cruciate ligament, MCL = medial cruciate ligament, MIA = monoiodoacetate, NR = not reported, NZW = New Zealand White, OARSI = osteoarthritis research society international, PCL = posterior cruciate ligament.

* = Lee *et al.* (2020): 11 animals (5 male and 4 female) were used across three groups: 2 OA groups and 1 control group.

| Study (year) | Country | Species (strain) | OA model | Tissue (details) | Stage of disease / OA severity | Control population | No. of animals | Female (%) | Age at start of study (w/m/y) | Follow-up | Ref |
|---|---|---|---|---|---|---|---|---|---|---|---|
| Almasry (2015) | Saudi Arabia | Rat (Wistar albino) | Meniscectomy | Synovium | Early stage | Non-operated | OA: 15<br>Control: 15 | 0<br>0 | Adult | 6w | [154] |
| Anderson-Mackenzie (1999) | UK | Mouse (CBA, STR/ort) | STR/ort | Ligament (ACL & PCL) | NR | CBA mice | OA: 16<br>Control: NR | 0<br>0 | 22/30/40/50w<br>22/30/40/50w | 0w | [137] |
| Bansal (2020) | USA | Pig (Yucutan minipig) | DMM | Meniscus | NR | Sham surgery | OA: 13<br>Control: 11 | 0<br>0 | 6.8m<br>6.8m | 1 & 3m | [152] |
| Barton (2021) | Canada | Sheep (Suffolk cross) | Partial ACL transection | Ligament (PCL), synovium | NR | Non-operated | OA: 11<br>Control: 6 | 100<br>100 | 3-5y<br>3-5y | 20 & 40 w | [141] |
| Bedingfield (2021) | USA | Mouse (C57BL/6) | Mechanical loading | Meniscus, synovium | DJD score 5 | Non-loaded | NR<br>NR | NR<br>NR | 6m<br>6m | NR<br>NR | [143] |
| Bryk (2021) | Poland | Rat (Wistar) | MIA | Synovium | NR | Non-injected | OA: 5-8 per time period<br>Control: 1-2 per group | 0<br>0 | NR<br>NR | 2-28d | [155] |
| Castrogiovanni (2019) | Italy | Rat (Wistar) | ACLT | Synovium | OARSI score | NR | OA: 8<br>Control: 8 | 0<br>0 | Adult<br>Adult | 14w | [156] |

| | | | | | | | | | | | |
|---|---|---|---|---|---|---|---|---|---|---|---|
| | | | | | 3.25 ± 0.71 | | | | | | |
| Dai (2020) | China | Rat (Sprague-Dawley) | ACLT | Synovium (ant. & post.) | Early to late-stage OA | Sham surgery | OA: 36<br>Control: 36 | 0<br>0 | 12w<br>12w | 4, 8 & 12w | [157] |
| Endo (2018) | Japan | Rat (Wistar) | ACLT | Meniscus (medial & lateral) | OARSI score 1 | Sham surgery | OA: 12<br>Control: 12 | 0<br>0 | 10w<br>10w | 3w | [151] |
| Funakoshi (2007) | Canada | Sheep (Suffolk cross) | ACL + MCL transection | Ligament (ACL & PCL) | NR | Non-operated | OA: 5<br>Control: 4 | 100<br>100 | 3y<br>3y | 20w | [142] |
| Gamal (2019) | Egypt | Rat (albino) | Cartilage defect | Synovium | NR | Non-operated | OA: 10<br>Control: 10 | 0<br>0 | 4-5m<br>4-5m | 4 & 6w | [158] |
| Hellio Le Graverand (2001) | France | Rabbit (NZW) | ACLT | Meniscus (medial & lateral) | NR | Non-operated | OA: 24<br>Control: 12 | 100<br>100 | 12m<br>12m | 3 & 8w | [146] |
| Hellio Le Graverand (2001) | Canada | Rabbit (NZW) | ACLT | Meniscus (medial) | NR | Non-operated | OA: 16<br>Control: 16 | 100<br>100 | 12m<br>12m | 1, 2, 3, 4w | [147] |
| Lapadula (1995) | Italy | Rabbit (Flanders giant) | Vitamin A injection | Synovium | Early stage | Untreated | OA: 12<br>Control: 4 | 0<br>0 | 8m<br>8m | 3, 6, 9d | [164] |
| Lee (2020) | USA | Mouse (BALB/cByJ) | DMM | Meniscus | NR | Sham surgery | OA: unclear*<br>Ctrl: unclear* | *<br>* | 12w<br>12w | 5w | [144] |
| Levillain (2017) | France | Rabbit (NZW) | ACLT | Meniscus (medial) | Early stage | Non-operated | OA: 6<br>Control: 6 | 0<br>0 | 6m<br>6m | 8w | [149] |
| Levillain (2017) | France | Rabbit (NZW) | ACLT | Meniscus (medial) | Early stage | Non-operated | OA: 6<br>Control: 6 | 0<br>0 | 6m<br>6m | 8w | [148] |
| Li (2020) | China | Rat (Sprague-Dawley) | MIA | Synovium | NR | Sham surgery | OA: 6<br>Control: 6 | 0<br>0 | 8w<br>8w | 8w | [159] |
| Loeser (2013) | USA | Mouse (C57BL/6) | DMM | Meniscus, ligament, capsule, synovium | Early to end-stage | Sham surgery | OA: 36<br>Control: 36 | 0<br>0 | 12w<br>12w | 2, 4, 8, 16w | [136] |

| | | | | | | | | | | | |
|---|---|---|---|---|---|---|---|---|---|---|---|
| McErlain (2008) | Canada | Rat (Sprague-Dawley) | ACLT | Tendon (patellar) | NR | Sham surgery | OA: 15<br>Control: 3 | 0<br>0 | 3w<br>3w | 1, 2, 3, 4, 5m | [166] |
| Miller (2014) | Canada | Rabbit (NZW) | ACLT | Ligament (MCL) | NR | Non-operated | OA: 6<br>Control: 6 | 100<br>100 | 1y<br>1y | 6w | [140] |
| Muschter (2020) | Germany | Mouse (C57BL/6) | DMM | Meniscus | NR | Sham surgery | OA: 72<br>Control: 72 | 0<br>0 | 8-10w<br>8-10w | 2, 4, 8, 12w | [145] |
| Ramos-Mucci (2020) | UK | Mouse (CBA, STR/ort) | STR/ort | Meniscus, ligaments (ACL, MCL, LCL) | OARSI grade 1-6 | CBA mice | OA: 29<br>Control: 12 | 0<br>0 | 27, 37, 40w<br>26, 40w | N/A | [138] |
| Shi (2020) | China | Rabbit (NZW) | Adapted Videman method | Skeletal muscle (rec. fem & biceps fem) | NR | Non-operated | OA: 6<br>Control: 6 | 0<br>0 | NR<br>NR | 10w | [153] |
| Walton (1977) | UK | Mouse (CBA, STR/ort) | STR/ort | Ligament | NR | CBA mice | OA: 351<br>Control: 348 | 36<br>28 | 4+m<br>4+m | NR<br>NR | [139] |
| Wei (2021) | China | Rabbit (domestic) | Modified Hulth method | Synovium | OARSI score ~5 (4w) to ~11 (12w) | Sham surgery | OA: 15<br>Control: 5 | NR<br>NR | 6m<br>6m | 4, 12w | [165] |
| Zhang (2021) | China | Rabbit (Sprague-Dawley) | ACLT, DMM, MIA | Synovium | NR | 'Normal' | Total: 80 | 0 | NR | 2, 4w | [161] |
| Zhang (2021) | China | Rabbit (Sprague-Dawley) | MIA | Synovium | NR | 'Normal' | OA: 8<br>Control: 8 | 0<br>0 | 2m<br>2m | 5w | [160] |
| Zhang (2019) | China | Rabbit (Sprague-Dawley) | MIA | Synovium | NR | Saline injection | OA: 6<br>Control: 6 | 100<br>100 | 3m<br>3m | 4w | [163] |
| Zhang (2019) | China | Rabbit (Sprague-Dawley) | ACLT | Synovium | NR | 'Normal' | OA: 8<br>Control: 8 | 0<br>0 | 3m<br>3m | 4w | [162] |

| | | | | | | | | | | | |
|---|---|---|---|---|---|---|---|---|---|---|---|
| Zhao (2014) | China | Rabbit<br>(Chinese) | Cartilage injury | Meniscus<br>(interior) | NR | Sham surgery | OA: 12<br>Control: 12 | 100<br>100 | 20m<br>20m | 1-6w | [150] |

**Supplementary Table 3**. 2015 OHAT risk of bias analysis of all included studies. Bias in each category is rated according to the following 4-point scale: ++, definitely low risk of bias; +, probably low risk of bias; -, probably high risk of bias; --, definitely high risk of bias. Questions 1-6 only apply to certain study types. Of note, several studies on human OA did not recruit a healthy control group, but instead performed subgroup analysis of a single cohort. In these cases, confounding bias was assessed in question 11.

Abbreviations: A/EB = attrition/exclusion bias, CB = confounding bias, DB = detection bias, N/A = not applicable, OB = other bias, PB = performance bias, SB = selection bias, SRB = selective reporting bias.

| **Domains** | **SB** | | | **CB** | **PB** | | **A/EB** | **DB** | | **SRB** | **OB** |
|---|---|---|---|---|---|---|---|---|---|---|---|
| **OHAT questions** | **1** | **2** | **3** | **4** | **5** | **6** | **7** | **8** | **9** | **10** | **11** |
| Abdul Sahib (2017) | N/A | N/A | - | - | N/A | N/A | + | - | -- | - | ++ |
| Abraham (2014) | N/A | N/A | - | - | N/A | N/A | ++ | - | + | ++ | ++ |
| Akisue (2002) | N/A | N/A | - | - | N/A | N/A | ++ | ++ | + | ++ | ++ |
| Allain (2001) | N/A | N/A | N/A | N/A | N/A | N/A | ++ | - | - | ++ | - |
| Almasry (2015) | + | - | N/A | N/A | -- | - | - | ++ | + | ++ | ++ |
| Anderson-Mackenzie (1999) | N/A | N/A | ++ | ++ | N/A | N/A | - | ++ | + | ++ | ++ |
| Bansal (2020) | - | - | N/A | N/A | + | - | - | ++ | ++ | ++ | ++ |
| Barton (2021) | - | - | N/A | N/A | - | - | - | + | ++ | + | ++ |
| Battistelli (2019) | N/A | N/A | - | - | N/A | N/A | - | ++ | - | -- | -- |
| Bedingfield (2021) | -- | -- | N/A | N/A | -- | -- | - | ++ | + | ++ | ++ |
| Belluzzi (2020) | N/A | N/A | ++ | ++ | N/A | N/A | - | - | + | ++ | ++ |
| Bryk (2021) | - | - | N/A | N/A | - | - | - | + | + | - | ++ |
| Cameron (1973) | N/A | N/A | - | - | N/A | N/A | + | - | - | - | - |
| Campbell (2015) | N/A | N/A | ++ | ++ | N/A | N/A | - | ++ | + | ++ | ++ |
| Castrogiovanni (2019) | - | - | N/A | N/A | - | - | - | ++ | ++ | ++ | ++ |
| Chang (2005) | N/A | N/A | - | - | N/A | N/A | - | - | + | ++ | ++ |
| Cheng (1996) | N/A | N/A | -- | ++ | N/A | N/A | ++ | ++ | ++ | ++ | ++ |
| Christensen (2019) | N/A | N/A | -- | -- | N/A | N/A | + | + | - | ++ | ++ |
| Cillero-Pastor (2015) | N/A | N/A | - | - | N/A | N/A | + | - | + | ++ | ++ |

| | | | | | | | | | | | |
|---|---|---|---|---|---|---|---|---|---|---|---|
| Cutolo (1992) | N/A | N/A | - | - | N/A | N/A | + | ++ | - | - | - |
| Dai (2020) | + | - | N/A | N/A | ++ | - | + | ++ | + | ++ | ++ |
| Dessombz (2013) | N/A | N/A | N/A | N/A | N/A | N/A | -- | ++ | + | + | ++ |
| DiCesare (1999) | N/A | N/A | - | - | N/A | N/A | + | - | + | + | ++ |
| DiFrancesco (1995) | N/A | N/A | + | + | N/A | N/A | ++ | - | - | - | + |
| Dijkgraaf (1997) | N/A | N/A | - | + | N/A | N/A | - | ++ | - | ++ | - |
| Doerschuk (1999) | N/A | N/A | - | - | N/A | N/A | - | ++ | - | + | ++ |
| Ea (2013) | N/A | N/A | - | - | N/A | N/A | + | ++ | ++ | ++ | - |
| Endo (2018) | + | - | N/A | N/A | + | - | - | ++ | - | + | ++ |
| Ene (2015) | N/A | N/A | - | - | N/A | N/A | + | + | - | -- | ++ |
| Exposito Molinero (2016) | N/A | N/A | + | + | N/A | N/A | + | - | + | ++ | ++ |
| Fan (2012) | N/A | N/A | + | + | N/A | N/A | ++ | ++ | - | - | ++ |
| Fink (2007) | N/A | N/A | N/A | N/A | N/A | N/A | + | ++ | - | ++ | ++ |
| Fischenich (2015) | N/A | N/A | - | - | N/A | N/A | - | - | ++ | ++ | ++ |
| Folkesson (2020) | N/A | N/A | ++ | ++ | N/A | N/A | ++ | ++ | + | ++ | ++ |
| Fuhrmann (2015) | N/A | N/A | + | + | N/A | N/A | ++ | - | ++ | ++ | - |
| Funakoshi (2007) | - | - | N/A | N/A | - | - | + | ++ | + | ++ | ++ |
| Gamal (2019) | - | - | N/A | N/A | - | - | + | + | - | ++ | ++ |
| Ghosh (1975) | N/A | N/A | - | - | N/A | N/A | - | + | + | - | ++ |
| Grevenstein (2020) | N/A | N/A | - | - | N/A | N/A | ++ | - | - | ++ | ++ |
| Haut Donahue (2021) | N/A | N/A | ++ | ++ | N/A | N/A | + | - | + | ++ | ++ |
| Heinegård (1968) | N/A | N/A | - | - | N/A | N/A | ++ | + | + | ++ | -- |
| LeGraverand (2001) (ref [146]) | - | - | N/A | N/A | - | - | - | + | + | ++ | ++ |
| LeGraverand (2001) (ref [147]) | - | - | N/A | N/A | - | - | - | + | - | ++ | ++ |
| Herbert (1973) | N/A | N/A | - | - | N/A | N/A | ++ | - | + | ++ | -- |
| Hino (1995) | N/A | N/A | - | - | N/A | N/A | + | + | - | -- | + |
| Hino (2020) | N/A | N/A | N/A | N/A | N/A | N/A | + | - | ++ | ++ | ++ |
| Ibrahim (2019) | N/A | N/A | ++ | ++ | N/A | N/A | + | + | + | ++ | ++ |
| Ibrahim (2021) | N/A | N/A | + | + | N/A | N/A | - | - | ++ | ++ | ++ |
| Ishizuka (2016) | N/A | N/A | -- | -- | N/A | N/A | -- | ++ | - | ++ | - |
| Itokazu (1998) | N/A | N/A | - | - | N/A | N/A | ++ | - | -- | ++ | -- |
| Jacquet (2018) | N/A | N/A | N/A | N/A | N/A | N/A | + | ++ | ++ | ++ | ++ |
| Jacquet (2019) | N/A | N/A | N/A | N/A | N/A | N/A | + | ++ | + | ++ | ++ |
| Johnson (2001) | N/A | N/A | - | - | N/A | N/A | - | - | - | ++ | - |
| Karjalainen (2021) | N/A | N/A | -- | ++ | N/A | N/A | ++ | ++ | + | ++ | ++ |

| | | | | | | | | | | | |
|---|---|---|---|---|---|---|---|---|---|---|---|
| Karube (1981) | N/A | N/A | - | - | N/A | N/A | ++ | - | + | ++ | - |
| Katsuragawa (2010) | N/A | N/A | + | + | N/A | N/A | - | ++ | - | ++ | ++ |
| Kaufmann (2003) | N/A | N/A | -- | -- | N/A | N/A | ++ | + | + | ++ | -- |
| Kiraly (2017) | N/A | N/A | N/A | N/A | N/A | N/A | + | - | - | - | ++ |
| Klareskog (1986) | N/A | N/A | - | - | N/A | N/A | + | - | - | - | ++ |
| Kodama (2018) | N/A | N/A | N/A | N/A | N/A | N/A | -- | ++ | - | ++ | ++ |
| Komro (2020) | N/A | N/A | -- | -- | N/A | N/A | + | ++ | + | ++ | + |
| Konttinen (1999) | N/A | N/A | - | - | N/A | N/A | ++ | - | - | ++ | - |
| Konttinen (2001) | N/A | N/A | + | + | N/A | N/A | - | - | + | ++ | ++ |
| Kragstrup (2019) | N/A | N/A | N/A | N/A | N/A | N/A | + | ++ | + | ++ | ++ |
| Kriegsmann (2004) | N/A | N/A | - | - | N/A | N/A | + | - | + | - | - |
| Kumagai (2012) | N/A | N/A | N/A | N/A | N/A | N/A | + | ++ | + | ++ | - |
| Kwok (2014) | N/A | N/A | - | - | N/A | N/A | ++ | ++ | - | ++ | - |
| Lapadula (1995) | + | - | N/A | N/A | - | - | + | - | - | - | -- |
| Lee (2020) | - | - | N/A | N/A | - | - | - | + | - | - | ++ |
| Levillain (2017) (ref [149]) | - | - | N/A | N/A | - | - | ++ | ++ | - | ++ | ++ |
| Levillain (2017) (ref [148]) | + | - | N/A | N/A | - | - | - | ++ | - | ++ | ++ |
| Levy (2013) | N/A | N/A | N/A | N/A | N/A | N/A | - | + | ++ | ++ | - |
| Li (2000) | N/A | N/A | - | - | N/A | N/A | + | - | - | ++ | - |
| Li (2020) | + | - | N/A | N/A | + | - | - | + | + | ++ | ++ |
| Loeser (2013) | + | - | N/A | N/A | ++ | - | + | ++ | + | ++ | ++ |
| Lopez-Franco (2016) | N/A | N/A | - | - | N/A | N/A | ++ | + | ++ | ++ | ++ |
| Mapp (1985) | N/A | N/A | - | - | N/A | N/A | + | - | + | ++ | - |
| Marczak (2017) | N/A | N/A | ++ | ++ | N/A | N/A | ++ | ++ | - | ++ | ++ |
| Martins (2018) | N/A | N/A | - | - | N/A | N/A | ++ | ++ | + | ++ | ++ |
| Masuda (1991) | N/A | N/A | + | + | N/A | N/A | + | -- | - | ++ | - |
| Mattiello-Svetzut (2013) | N/A | N/A | N/A | N/A | N/A | N/A | -- | ++ | ++ | ++ | ++ |
| Mazzocca (2013) | N/A | N/A | + | + | N/A | N/A | -- | + | ++ | ++ | ++ |
| McDaniel (2017) | N/A | N/A | ++ | ++ | N/A | N/A | + | ++ | - | ++ | ++ |
| McErlain (2008) | + | - | N/A | N/A | ++ | - | ++ | ++ | - | ++ | ++ |
| Meknas (2012) | N/A | N/A | - | - | N/A | N/A | + | - | + | ++ | ++ |
| Melrose (2008) | N/A | N/A | ++ | ++ | N/A | N/A | - | - | - | ++ | - |
| Miller (2014) | - | ++ | N/A | N/A | ++ | ++ | ++ | - | ++ | ++ | -- |
| Mine (2013) | N/A | N/A | - | - | N/A | N/A | - | - | - | ++ | - |
| Mobargha (2014) | N/A | N/A | N/A | N/A | N/A | N/A | + | ++ | - | ++ | ++ |

| | | | | | | | | | | | |
|---|---|---|---|---|---|---|---|---|---|---|---|
| Monibi (2021) | N/A | N/A | - | - | N/A | N/A | - | - | ++ | + | ++ |
| Muschter (2020) | - | - | N/A | N/A | ++ | - | -- | ++ | ++ | ++ | ++ |
| Musumeci (2014) | N/A | N/A | + | + | N/A | N/A | + | ++ | ++ | ++ | ++ |
| Nagata (2000) | N/A | N/A | - | - | N/A | N/A | ++ | ++ | - | ++ | - |
| Nakahara (2013) | N/A | N/A | + | + | N/A | N/A | -- | ++ | - | -- | + |
| Nakashima (1998) | N/A | N/A | N/A | N/A | N/A | N/A | ++ | ++ | - | ++ | ++ |
| Nelissen (2001) | N/A | N/A | - | - | N/A | N/A | ++ | ++ | - | ++ | - |
| Nikkari (1995) | N/A | N/A | - | - | N/A | N/A | ++ | - | - | ++ | - |
| Nishida (1995) | N/A | N/A | - | - | N/A | N/A | + | - | - | ++ | - |
| Noehren (2018) | N/A | N/A | + | + | N/A | N/A | - | ++ | ++ | ++ | ++ |
| Okamoto (2015) | N/A | N/A | + | + | N/A | N/A | ++ | ++ | - | ++ | - |
| Park (2015) | N/A | N/A | - | - | N/A | N/A | + | ++ | ++ | ++ | ++ |
| Park (2021) | N/A | N/A | N/A | N/A | N/A | N/A | ++ | - | + | ++ | ++ |
| Poduval (2010) | N/A | N/A | - | - | N/A | N/A | - | - | - | ++ | - |
| Pollock (1990) | N/A | N/A | - | - | N/A | N/A | + | - | - | ++ | - |
| Pordzik (2020) | N/A | N/A | + | + | N/A | N/A | + | ++ | + | ++ | ++ |
| Rafael (2014) | N/A | N/A | - | - | N/A | N/A | - | - | - | ++ | - |
| Rajgopal (2014) | N/A | N/A | N/A | N/A | N/A | N/A | ++ | + | + | ++ | ++ |
| Ramos-Mucci (2020) | N/A | N/A | ++ | ++ | N/A | N/A | - | ++ | - | - | - |
| Richardot (2009) | N/A | N/A | - | - | N/A | N/A | ++ | ++ | - | ++ | ++ |
| Rinaldi (1998) | N/A | N/A | - | - | N/A | N/A | - | ++ | + | ++ | - |
| Roller (2015) (ref [78]) | N/A | N/A | ++ | ++ | N/A | N/A | + | ++ | + | ++ | ++ |
| Roller (2015) (ref [79]) | N/A | N/A | - | - | N/A | N/A | -- | ++ | ++ | + | - |
| Saito (2002) | N/A | N/A | N/A | N/A | N/A | N/A | - | ++ | + | ++ | ++ |
| Santiago (2006) | | | | | | | | | | | |
| Schneider (1994) | N/A | N/A | - | - | N/A | N/A | + | + | - | ++ | - |
| Scott (1981) | N/A | N/A | - | - | N/A | N/A | + | - | - | ++ | - |
| Scott (1984) | N/A | N/A | - | - | N/A | N/A | ++ | - | - | ++ | - |
| Serrão (2014) | N/A | N/A | ++ | ++ | N/A | N/A | ++ | ++ | + | ++ | - |
| Shi (2020) | + | - | N/A | N/A | + | - | - | + | + | + | ++ |
| Sladojevic (2016) | N/A | N/A | - | - | N/A | N/A | + | - | + | ++ | ++ |
| Son (2013) | N/A | N/A | N/A | N/A | N/A | N/A | + | - | + | ++ | ++ |
| Sun (2010) | N/A | N/A | - | - | N/A | N/A | + | - | ++ | ++ | ++ |
| Sun (2012) | N/A | N/A | - | - | N/A | N/A | ++ | - | ++ | ++ | ++ |
| Takahashi (1996) | N/A | N/A | - | - | N/A | N/A | + | ++ | + | ++ | ++ |
| Takahashi (1998) | N/A | N/A | -- | ++ | N/A | N/A | + | + | + | - | ++ |

| | | | | | | | | | | | |
|---|---|---|---|---|---|---|---|---|---|---|---|
| Turdean (2017) | N/A | N/A | - | - | N/A | N/A | ++ | ++ | + | - | -- |
| Van Linthoudt (1997) | N/A | N/A | N/A | N/A | N/A | N/A | ++ | ++ | + | ++ | ++ |
| Walton (1977) | N/A | N/A | - | - | N/A | N/A | - | + | - | - | + |
| Wang (2018) | N/A | N/A | - | - | N/A | N/A | + | ++ | + | ++ | ++ |
| Wang (2020) | N/A | N/A | N/A | N/A | N/A | N/A | ++ | ++ | ++ | ++ | ++ |
| Warnecke (2020) | N/A | N/A | - | - | N/A | N/A | - | - | + | ++ | ++ |
| Wei (2021) | + | - | N/A | N/A | - | - | + | ++ | ++ | ++ | ++ |
| Worrall (1991) | N/A | N/A | - | - | N/A | N/A | + | ++ | - | - | - |
| Worrall (1994) | N/A | N/A | - | - | N/A | N/A | + | ++ | - | ++ | ++ |
| Zhang (2012) | N/A | N/A | N/A | N/A | N/A | N/A | ++ | - | + | ++ | - |
| Zhang (2019) ref ([163]) | + | - | N/A | N/A | ++ | - | + | + | - | - | ++ |
| Zhang (2019) ref ([162]) | + | - | N/A | N/A | - | - | - | + | - | - | ++ |
| Zhang (2021) ref ([161]) | ++ | - | N/A | N/A | - | - | + | + | + | + | ++ |
| Zhang (2021) ref ([160]) | + | - | N/A | N/A | - | - | + | + | + | ++ | ++ |
| Zhao (2014) | + | - | N/A | N/A | ++ | - | + | ++ | - | ++ | - |
| Zhu (2012) | N/A | N/A | -- | -- | N/A | N/A | - | - | + | ++ | - |

**Supplementary Information – search strategy**

**Database: Medline (Ovid MEDLINE® Epub Ahead of Print, In-Process & Other Non-Indexed Citations, Ovid MEDLINE® Daily and Ovid MEDLINE®) 1946 to present**

Search Strategy:

--------------------------------------------------------------------------------

1 exp Proteoglycans/ (39384)

2 exp Elasticity/ (50892)

3 (elasticity or "elastic modulus" or topograph* or anisotrop*).ti,ab. (160630)

4 exp Extracellular Matrix/ (47818)

5 "extracellular matri*".ti,ab. (106140)

6 exp Extracellular Matrix Proteins/ (193763)

7 (Aggrecan* or "Cartilage Specific Proteoglycan Core Proteins" or COMP or "Thrombospondin 5" or TSP5 or collagen* or pro-collagen* or procollagen or tropocollagen* or fibulin* or "alpha-collagen" or elastin* or thropoelastin* or fibrillin* or fibronectin* or glycoprotein* or laminin* or "glycoprotein GP-2" or "latent TGF beta binding protein" or matrilin* or MATN* or "matrix protein*" or netrin* or netrin-1 or biglycan* or decorin* or hyalectin* or neurocan* or fibromodulin* or lumican* or tenacin* or tenacin-c or cytotactin* or hexabrachion* or versican* or "chondroitin sulfate proteoglycan core protein 2" or "chondroitin sulphate proteoglycan core protein 2" or vitronectin* or pyridinoline* or hydroxylysylpyridinoline*).ti,ab. (448428)

8 microfibril*.ti,ab. (4325)

9 exp Glycosaminoglycans/ (121397)

10 (glycosaminoglycan* or mucopolysaccharide* or proteoglycan* or chondroitin or translagen or blutal or heparin or "heparinic acid" or heparitin or heparan or "hyaluronic acid" or hyaluron*n or hyaluronate or keratan or keratosulphate or keratosulfate).ti,ab. (160866)

11 Calcinosis/ (38846)

12 (calcinosis or calcinoses or calcification* or microcalcification* or microcalcinosis or microcalcinoses or hydroxyapatite or hydroxylapatite).ti,ab. (93466)

13 1 or 2 or 3 or 4 or 5 or 6 or 7 or 8 or 9 or 10 or 11 or 12 (1009050)

14 exp Intervertebral Disc/ (15222)

15 ("intervertebral dis*" or "annulus fibrosus" or "anulus fibrosis" or "nucleus pulposus").ti,ab. (18128)

16 exp Ligaments, Articular/ (32725)

17 ligament*.ti,ab. (85003)

18 exp Muscle, Skeletal/ (283110)

19 muscle*.ti,ab. (744181)

20 exp Tendons/ (44653)

21 tendon*.ti,ab. (70548)

22 Menisci, Tibial/ (7362)

23 (menisc* or "semilunar cartilage*").ti,ab. (18889)

24 exp Joint Capsule/ (29528)
25 ("joint capsule*" or "articular capsule*" or "capsula articularis" or "synovial capsule*" or "synovial fold*" or "synovial plica*" or "hip capsule*" or "knee capsule*" or "shoulder capsule*").ti,ab. (3859)
26 Bursa, Synovial/ (1124)
27 ("synovial bursa*" or synovium or "synovial membrane*").ti,ab. (11261)
28 exp Adipose Tissue/ (103687)
29 ("adipose tissue*" or "body fat*" or "body pad*" or "fat* tissue*" or "fat pad*").ti,ab. (123620)
30 14 or 15 or 16 or 17 or 18 or 19 or 20 or 21 or 22 or 23 or 24 or 25 or 26 or 27 or 28 or 29 (1173134)
31 exp Osteoarthritis/ (68777)
32 (osteoarthr* or "degenerative arthri*" or arthroses or arthrosis).ti,ab. (85531)
33 31 or 32 (105393)
34 13 and 30 and 33 (4268)
35 34 (4268)
36 limit 35 to yr="2020 - 2021" (490)

**Database: Embase 1974 to present**

Search Strategy:

--------------------------------------------------------------------------------

1 proteoglycan/ (24376)
2 exp elasticity/ (69514)
3 (elasticity or "elastic modulus" or topograph* or anisotrop*).ti,ab. (166957)
4 exp extracellular matrix/ (153794)
5 "extracellular matri*".ti,ab. (134542)
6 exp scleroprotein/ (346147)
7 (Aggrecan* or "Cartilage Specific Proteoglycan Core Proteins" or COMP or "Thrombospondin 5" or TSP5 or collagen* or pro-collagen* or procollagen or tropocollagen* or fibulin* or "alpha-collagen" or elastin* or thropoelastin* or fibrillin* or fibronectin* or glycoprotein* or laminin* or "glycoprotein GP-2" or "latent TGF beta binding protein" or matrilin* or MATN* or "matrix protein*" or netrin* or netrin-1 or biglycan* or decorin* or hyalectin* or neurocan* or fibromodulin* or lumican* or tenacin* or tenacin-c or cytotactin* or hexabrachion* or versican* or "chondroitin sulfate proteoglycan core protein 2" or "chondroitin sulphate proteoglycan core protein 2" or vitronectin* or pyridinoline* or hydroxylysylpyridinoline*).ti,ab. (555848)
8 microfibril*.ti,ab. (4450)
9 exp glycosaminoglycan/ (250102)
10 (glycosaminoglycan* or mucopolysaccharide* or proteoglycan* or chondroitin or translagen or blutal or heparin or "heparinic acid" or heparitin or heparan or "hyaluronic acid" or hyaluron*n or hyaluronate or keratan or keratosulphate or keratosulfate).ti,ab. (203041)
11 exp calcinosis/ (16203)

12 (calcinosis or calcinoses or calcification* or microcalcification* or microcalcinosis or microcalcinoses or hydroxyapatite or hydroxylapatite).ti,ab. (118865)
13 1 or 2 or 3 or 4 or 5 or 6 or 7 or 8 or 9 or 10 or 11 or 12 (1369541)
14 exp intervertebral disk/ (16400)
15 ("intervertebral dis*" or "annulus fibrosus" or "anulus fibrosis" or "nucleus pulposus").ti,ab. (21310)
16 exp joint ligament/ (28620)
17 ligament*.ti,ab. (103603)
18 exp skeletal muscle/ (367283)
19 muscle*.ti,ab. (919762)
20 exp tendon/ (41624)
21 tendon*.ti,ab. (82988)
22 knee meniscus/ (10357)
23 (menisc* or "semilunar cartilage*").ti,ab. (22980)
24 joint capsule/ (3803)
25 ("joint capsule*" or "articular capsule*" or "capsula articularis" or "synovial capsule*" or "synovial fold*" or "synovial plica*" or "hip capsule*" or "knee capsule*" or "shoulder capsule*").ti,ab. (4819)
26 synovial bursa/ (2550)
27 ("synovial bursa*" or synovium or "synovial membrane*").ti,ab. (15675)
28 exp adipose tissue/ (176340)
29 ("adipose tissue*" or "body fat*" or "body pad*" or "fat* tissue*" or "fat pad*").ti,ab. (166084)
30 14 or 15 or 16 or 17 or 18 or 19 or 20 or 21 or 22 or 23 or 24 or 25 or 26 or 27 or 28 or 29 (1411380)
31 exp osteoporosis/ (138070)
32 (osteoarthr* or "degenerative arthri*" or arthroses or arthrosis).ti,ab. (119642)
33 31 or 32 (253893)
34 13 and 30 and 33 (6543)
35 34 (6543)
36 limit 35 to yr="2020 - 2021" (909)

**Scopus**

( ( TITLE-ABS-KEY ( elasticity OR "elastic modulus" OR topograph* OR anisotrop* OR "extracellular matri*" OR aggrecan* OR "Cartilage Specific Proteoglycan Core Proteins" OR comp OR "Thrombospondin 5" OR tsp5 OR collagen* OR pro-collagen* OR procollagen OR tropocollagen* OR fibulin* OR "alpha-collagen" OR elastin* OR thropoelastin* OR fibrillin* OR fibronectin* OR glycoprotein* OR laminin* OR "glycoprotein GP-2" OR "latent TGF beta binding protein" OR matrilin* OR matn* OR "matrix protein*" OR netrin* OR netrin-1 OR biglycan* OR decorin* OR hyalectin* OR neurocan* OR fibromodulin* OR lumican* OR tenacin* OR tenacin-c OR cytotactin* OR hexabrachion* OR versican* OR "chondroitin sulfate proteoglycan core protein 2" OR "chondroitin sulphate proteoglycan core protein 2" OR vitronectin* OR pyridinoline* OR hydroxylysylpyridinoline* ) OR TITLE-ABS-KEY ( microfibril* OR glycosaminoglycan* OR mucopolysaccharide* OR proteoglycan* OR chondroitin OR translagen OR blutal OR heparin OR "heparinic acid" OR heparitin OR heparan OR "hyaluronic acid" OR hyaluron*n OR hyaluronate OR keratan OR keratosulphate OR keratosulfate OR calcinosis OR calcinoses OR calcification* OR microcalcification* OR microcalcinosis OR microcalcinoses OR hydroxyapatite OR hydroxylapatite ) ) ) AND ( ( TITLE-ABS-KEY ( ( "intervertebral dis*" OR "annulus fibrosus" OR "anulus fibrosis" OR "nucleus pulposus" ) ) OR TITLE-ABS-KEY ( ligament* OR muscle* OR tendon* OR menisc* OR "semilunar cartilage*" OR ( "joint capsule*" OR "articular capsule*" OR "capsula articularis" OR "synovial capsule*" OR "synovial fold*" OR "synovial plica*" OR "hip capsule*" OR "knee capsule*" OR "shoulder capsule*" ) OR ( "synovial bursa*" OR synovium OR "synovial membrane*" ) OR ( "adipose tissue*" OR "body fat*" OR "body pad*" OR "fat* tissue*" OR "fat pad*" ) ) ) ) AND ( TITLE-ABS-KEY ( osteoarthr* OR "degenerative arthri*" OR arthroses OR arthrosis ) ) AND ( LIMIT-TO ( PUBYEAR , 2022 ) OR LIMIT-TO ( PUBYEAR , 2021 ) OR LIMIT-TO ( PUBYEAR , 2020 ) )
